# Cumulative Indicators of Circadian Rhythm Disturbance and its Clinical, Functional and Genetic Correlates in the Brisbane Longitudinal Twin Study

**DOI:** 10.64898/2026.09.13.26362400

**Authors:** Mirim Shin, Emiliana Tonini, Joanne S. Carpenter, Brittany L. Michell, Baptiste Couvy-Duchesne, Shin Ho Park, Richard Parker, Enda M. Byrne, Samuel J. Hockey, Anna Treneman, Timothy To, Nayonika Bhattacharya, Andrew Shim, Elizabeth M. Scott, Nathan A. Gillespie, Nicholas G. Martin, Jan Scott, Sarah E. Medland, Jacob J. Crouse, Ian B. Hickie

## Abstract

**Background:** Circadian rhythm disturbances (CRDs) are transdiagnostic risk factors across mental disorders. We examined CRD-associated phenotypes, their heritability, and clinical, functional and genetic correlates, cross-sectionally, in young adults from the Brisbane Longitudinal Twin Study.

**Methods:** We assessed 2,773 community-based participants (25.6±4.1 years; 57.8% female). Six CRD-associated phenotypes (seasonality, hypersomnia, social jetlag, delayed sleep, evening preference, sleep inertia) were included. Associations of CRD-load (number of CRDs) with clinical and functional outcomes, and with polygenic risk scores (PRS) for mental health, physical health, and sleep/circadian traits, were tested using linear, negative binomial, or logistic regression, with family-clustered standard errors. Heritability of CRD-phenotypes was estimated using structural equation modelling.

**Results:** Almost half the sample (49.2%) reported experiencing ≥1 CRD-phenotype and 20.5% reported ≥2. Four CRD-phenotypes (hypersomnia, social jetlag, delayed sleep, evening preference) were significantly heritable (h^2^=0.19-0.48). Higher CRD-load was associated with greater psychological distress (β=0.96, p<0.001), more psychological symptoms (incidence rate ratio [IRR]=1.07-1.40), higher odds of a full-threshold mental disorder (OR=1.28, p<0.001), and greater functional impairment (days-in-bed, IRR=1.55; days-out-of-role, IRR=1.43). In genotyped participants (n=2,301), CRD-load was associated with mood-disorder PRS (major depression [OR=1.14, p=0.003] and bipolar disorder [OR=1.14, p=0.002]) but not PRSs for schizophrenia, attention deficit hyperactivity disorder, autism, neuroticism or anxiety. Trends towards increased PRSs for metabolic traits did not survive Bonferroni-correction.

**Conclusions:** CRD-load was associated with clinical symptoms, functional impairment, and mood-related genetic liability. These findings support further evaluation of brief assessments of circadian-associated phenotypes for identifying young people at greater clinical burden and informing circadian-focused prevention and early intervention strategies.

## INTRODUCTION

Sleep and circadian rhythm (‘body clock’) disturbances are transdiagnostic risk factors for mental disorders, with bidirectional relationships reported across mood, psychotic, and other disorders (Balter et al. 2024; Harvey and Sarfan 2024; Hickie and Crouse 2024; Meyer et al. 2024). Although the direction of causation remains unresolved, circadian rhythm disturbance (CRD) may represent a shared biological pathway underlying vulnerability to mood disorders, particularly bipolar spectrum disorders and atypical depression, which are characterized by dysregulated activation and energy, early age-of-onset, and poor response to conventional treatments (Carpenter et al. 2021; de Haan et al. 2026; Hickie et al. 2019). Identifying CRD during adolescence and young adulthood, when normative circadian phase delay coincides with the peak age of mood disorder onset (Carpenter et al. 2021; Logan et al. 2018; Pifer et al. 2024), may provide opportunities for targeted early interventions.

Notably, longitudinal studies have shown that hypersomnia (Breslau et al. 1996) and an elevated cortisol awakening response (Adam et al. 2010) predict incident depression. Prospective cohort studies have linked evening chronotype to an increased incidence of major depressive disorder (MDD) and bipolar disorder (Burns et al. 2024). Mendelian randomization studies using genetic instruments for diurnal preference report concordant, though not uniform, effects on depression risk (Daghlas et al. 2021; O’Loughlin et al. 2021). Actigraphy studies from genetically-informative, population, and clinical samples have linked reduced rest-activity amplitude and greater sleep-wake variability to mood disorders (Chen et al. 2022; Esaki et al. 2021; Pagani et al. 2016). More recently, studies of young people with emerging mood disorders have identified altered 24-hour wrist skin temperature rhythms (Shin et al. 2025a), delayed rest-activity rhythms (Carpenter et al. 2025a), and misalignment between internal circadian phase markers (Carpenter et al. 2025b). Importantly, evidence suggests that composite measures incorporating multiple CRD-associated phenotypes predict the onset of major mental illness more strongly than individual markers alone (Scott et al. 2020).

Despite this accumulating evidence, several gaps remain. CRD is a multidimensional construct (Coelho et al. 2025), yet few studies have developed composite CRD measures suitable for population-level screening. Traditional circadian assessments (e.g., dim light melatonin onset [DLMO], core body temperature monitoring, and actigraphy) while essential for research, are often expensive, invasive, or impractical for large population studies or low-resource settings (Wellcome Trust 2022). Finally, the extent to which genetic liability for mental health, physical health and sleep/circadian traits is associated with CRD remains underexplored in young adult populations.

In this study, we operationalized CRD as a dimensional, transdiagnostic phenotype in a large community-based twin and sibling cohort of young adults (Brisbane Longitudinal Twin Study; BLTS). We examined cumulative CRD-load (the number of CRD phenotypes endorsed) using a pragmatic selection of six accessible, survey-based phenotypes aligned with our proposed construct of *circadian depression* (seasonality, hypersomnia, social jetlag, delayed sleep, evening preference, and sleep inertia) (Carpenter et al. 2021).

Here, we aimed to (1) characterize the prevalence of CRD phenotypes and their clinical and functional correlates; (2) estimate the heritability of CRD phenotypes; and (3) test associations between cumulative CRD-load and polygenic risk scores (PRS) for mental health, physical health, and sleep/circadian traits. Twin-model heritability estimated the overall contribution of additive genetic factors to CRD phenotypes, whereas PRS analyses tested whether CRD-load captured specific genetic liabilities across related domains. Analyses were conducted in the full sample, with secondary analyses in the clinical subsample (those with mental disorders) to test for amplification of association.

We hypothesized that greater CRD-load would be associated with more severe mental health symptoms and functional impairment, and with stronger genetic liability to mental disorders, particularly mood disorders (bipolar disorder and major depression), alongside physical health (e.g., type 2 diabetes, insulin resistance, inflammation) and circadian-associated traits (e.g., chronotype), but not other sleep traits (e.g., insomnia). For heritability, we expected moderate additive genetic influence on the sleep-timing measures.

## METHODS

### Study design and participants

This study analyzed data from the BLTS, a prospective community-based cohort of twins and their non-twin siblings recruited in the greater Brisbane-area (Australia) via media appeals and by word of mouth from 1992 at age ∼12, with follow-up assessments at ages 14, 16, 19, and 25 (Gillespie et al. 2013; Wright and Martin 2004). The current study focuses on the “Nineteen and Up” (19Up) study (2009-2016), the first wave using the Composite International Diagnostic Interview (CIDI) assessment of mental disorders. Ethical approval was obtained from the Human Research Ethics Committee at the Queensland Institute of Medical Research (EC00278, P1212), with written informed consent obtained from all participants and parents/carers of those under 18. The detailed cohort profile is available elsewhere (Couvy-Duchesne et al. 2018).

### Measures

#### Demographic and clinical measures

Age, sex, twin status, marital status, and educational attainment were collected using questionnaires. Self-report scales were used to assess mental health symptoms including the Somatic and Psychological Health Report (SPHERE-12) for somatic and psychological symptoms (Hickie et al. 2001); five hypomanic symptoms using a modified version of the Altman Self-Rating Mania (Altman et al. 1997); six psychotic-like symptoms using a tool adapted in part from the Community Assessment of Psychic Experiences (Konings et al. 2006); and psychological distress using the Kessler 6-item Psychological Distress Scale (K6) (Kessler et al. 2002). Self-rated functioning (days-out-of-role and days-in-bed in the past-month) was assessed using two modified items from the WHO Disability Assessment Schedule (Ustün et al. 2010). Migraine was identified by self-report or by International Classification of Headache Disorders criteria for migraine without aura (Headache Classification Committee of the International Headache Society 2013). The specific questions and response options are presented in the Supplementary Materials. The CIDI assessment (Kessler et al. 2005) using item-based criteria was used to determine the presence or absence of DSM-IV mental disorders and age-of-onset for depressive, hypomanic, manic, anxiety (agoraphobia, panic disorder, social anxiety), or psychotic syndromes (schizophreniform/other psychoses). Primary analyses included all participants; subgroup analyses focused on specific mental disorders are reported in the Supplementary Materials.

#### CRD phenotypes

Six survey-based CRD-associated phenotypes were selected based on their availability in the BLTS and alignment with our proposed criteria for “circadian depression” (Carpenter et al. 2021; Tonini et al. 2026). As these measures were assessed independently of current mood episodes, they were treated as trait-like indicators of CRD rather than state markers. Established thresholds were used where available (seasonality), whereas continuous measures were dichotomized at +1 standard deviation (SD) above the sample mean to indicate the presence/absence of CRD rather than gradations within the typical range. This threshold is pragmatic rather than diagnostically validated.

**1. Seasonality**: Global Seasonality Score (GSS) ≥11, indicating moderate seasonality (Rosenthal et al. 1984)
**2. Hypersomnia\***: Weekend sleep duration ≥10 hours
**3. Social Jetlag\***: Difference in sleep midpoint between free days (i.e., weekends) and workdays (i.e., weekday) ≥2.25 hours
**4. Delayed Sleep\***: Weekend sleep midpoint later than 5:15am
**5. Evening Sleep Preference\***: Midpoint of sleep on ideal days later than 3:45am
**6. Sleep inertia**: Responding “Very sleepy” to the question “Over the past week, how did you feel when you woke up?”

*Thresholds were set at +1 SD above the sample mean for the primary analyses; results using age-specific thresholds (18-24 vs ≥25 years) are reported in the Supplementary Materials.

Seasonality was assessed only during the computer-assisted telephone interview phase (2009-2011) and was not retained in the subsequent online surveys (2012-2016), which accounts for the smaller number of responses for this phenotype (n=625). *CRD-load*, a dimensional construct, was defined as the number of phenotypes met and calculated as an unweighted sum, because each phenotype captures a different aspect of CRD. CRD-load was calculated for participants responding to at least two phenotypes (n=2,651); those with fewer than two valid responses were excluded (n=122).

### Estimating Heritability of CRD Phenotypes

Twin-based heritability was estimated for the continuous variables underlying each CRD phenotype rather than for the dichotomized phenotypes, as dichotomization of continuous traits reduces statistical power in twin models. Seven continuous measures were modelled: weekend sleep duration, weekday sleep midpoint, weekend sleep midpoint, sleep midpoint on ideal days, social jetlag, seasonality (GSS), and the sleep-inertia item. These seven measures index six CRD phenotypes, as social jetlag is derived from weekday and weekend sleep midpoints. Weekday sleep midpoint was additionally included as a timing measure in the heritability analyses. Models were fitted using structural equation modelling in OpenMx (v2.22.11).

Prior to estimating heritability, a series of assumption checking analyses were run to check for differences in the means, variances and covariances relating to birth-order, zygosity and sex (Evans et al. 1999). Sex and age effects were included on the means. Univariate models were used to decompose phenotypic variance into additive genetic (A), unique environmental (E) components and shared environmental (C) or dominant genetic (D) effects. A sex-limitation framework was used to test qualitative (different genetic factors by sex) and quantitative (different parameter magnitudes by sex) sex differences in the variance components. The best-fitting model for each phenotype was selected based on likelihood ratio tests, the Akaike Information Criterion (AIC), and parsimony. Standardized variance components with 95% confidence intervals (CIs) are reported separately by sex where the best-fitting model retained sex differences.

### Polygenic risk scores (PRS)

DNA was obtained from blood samples collected at the 12- or 14-year clinic visits. Pre-imputation quality control was done using PLINK 1.9 (Chang et al. 2015; Purcell et al. 2007). Quality control involved removing SNPs with a minor allele frequency <0.005, a SNP call rate <97.5%, and Hardy-Weinberg equilibrium (p<1x10^-6^), before imputation using the HRC v1.1 reference panel (Taliun et al. 2021). Ancestry was inferred using ancestrally-informative genetic principal components (PCs) by projecting PCs on 1000 Genomes data. Analyses were restricted to participants of genetically inferred European ancestry, as nearly all participants were of European ancestry (Couvy-Duchesne et al. 2018). The latest publicly available genome-wide association study (GWAS) results for mental disorders, metabolic-inflammatory, and sleep/circadian traits (see Supplementary Materials) were used to calculate weights for PRS calculation. Where applicable, leave-one-out summary statistics were used for GWAS studies that included participants from the BLTS to avoid over-estimation. SBayesRC was used to generate allele weights for each PRS (Zheng et al. 2024). The posterior SNP effects for each disorder and trait were used to generate PRS for each participant using the PLINK score function (Purcell et al. 2007). Each PRS was standardized. Effect sizes are interpretable per SD unit increase.

### Statistical analysis

All statistical analyses were conducted using R version 4.5.0 (R Core Team 2025). Associations between CRD-load and clinical/functional outcomes were examined using regression models, adjusted for age, sex, and twin status. The eight primary outcomes were psychological distress (K6), hypomanic-like experiences, psychotic-like experiences, SPHERE-12 somatic symptoms and psychological symptoms, days-out-of-role, days-in-bed, and migraine. Meeting CIDI criteria for a full-threshold mental disorder was modelled separately, and age-of-onset was examined within the clinical subsample. Linear regression was used for continuous outcomes (K6 score). Negative binomial regression was used for count outcomes (hypomanic-like and psychotic-like experiences, SPHERE somatic and psychological symptoms, days-out-of-role, days-in-bed) showing overdispersion, skewness, or zero-inflation, based on variance-to-mean ratios >1 (range: 2.25-4.23). Logistic regression was used for migraine cases. Results are presented as unstandardized regression coefficients (β), incidence rate ratios (IRR) and odds ratios (OR), respectively. Multiple comparisons were Bonferroni-corrected across the eight primary outcomes (α=0.006).

PRSs were calculated for 28 traits across three domains: (i) seven mental health (e.g., ADHD, major depression, bipolar disorder, schizophrenia); (ii) 16 physical health (e.g., body mass index [BMI], insulin resistance, interleukin [IL]-1β/6/10); and (iii) five sleep/circadian (chronotype, sleep midpoint, insomnia, sleep duration, low relative amplitude [RA]). See Table S2 for the full list and GWAS sources. Associations between PRS and CRD-load were modelled using ordinal logistic regression with CRD-load as the outcome, adjusting for age, sex, twin status, and the first four ancestry principal components (PCs). Bonferroni correction was applied within each domain (mental health α=0.007; physical health α=0.003; sleep/circadian α=0.010). For all regression analyses (clinical correlate and PRS models), standard errors and 95% CIs were estimated using cluster-robust (sandwich) variance estimators clustered by family ID to account for the non-independence of related individuals (twins and non-twin siblings).

### Sensitivity Analyses

Four sensitivity analyses examined the robustness of CRD-load associations: (i) restricting analyses to complete responders (n=625) who provided valid responses to all six CRD phenotypes; (ii) restricting the CRD-load score to the phenotypes showing evidence of additive genetic influence in the heritability analyses; (iii) examining PRS associations separately in females and males; and (iv) repeating analyses after redefining each CRD phenotype using age-specific thresholds. Full results are reported in the Supplementary Materials (Tables S8-S12, Figures S1-S7) and in the Supplementary Data (SD1-SD19).

## RESULTS

### Participant characteristics and CRD phenotype distribution

Of 2,773 participants (mean age 25.6±4.1 years [range: 18-38]; 57.8% female), 45.1% were dizygotic twins, 44.7% had completed undergraduate degrees, 76.3% had never married and 84.8% had either work or study (Table 1). Overall, 36.0% (n=999) met CIDI criteria for mood, anxiety, or psychotic disorders. Of 2,651 participants with valid data for at least two CRD phenotypes, 1,305 (49.2%) met ≥1 phenotype and 543 (20.5%) met ≥2. The distribution was: one phenotype (n=762, 28.7%), two (n=383, 14.4%), three (n=137, 5.2%), four (n=19, 0.7%), and five (n=4, 0.2%); none met all six (Figure 1). Across all participants, the prevalence of individual CRD phenotypes was highest for hypersomnia (16.7%), followed by social jetlag (15.3%), sleep inertia (15.3%), delayed sleep (14.6%), evening preference (12.7%), and seasonality (2.2%).

**Figure 1.**
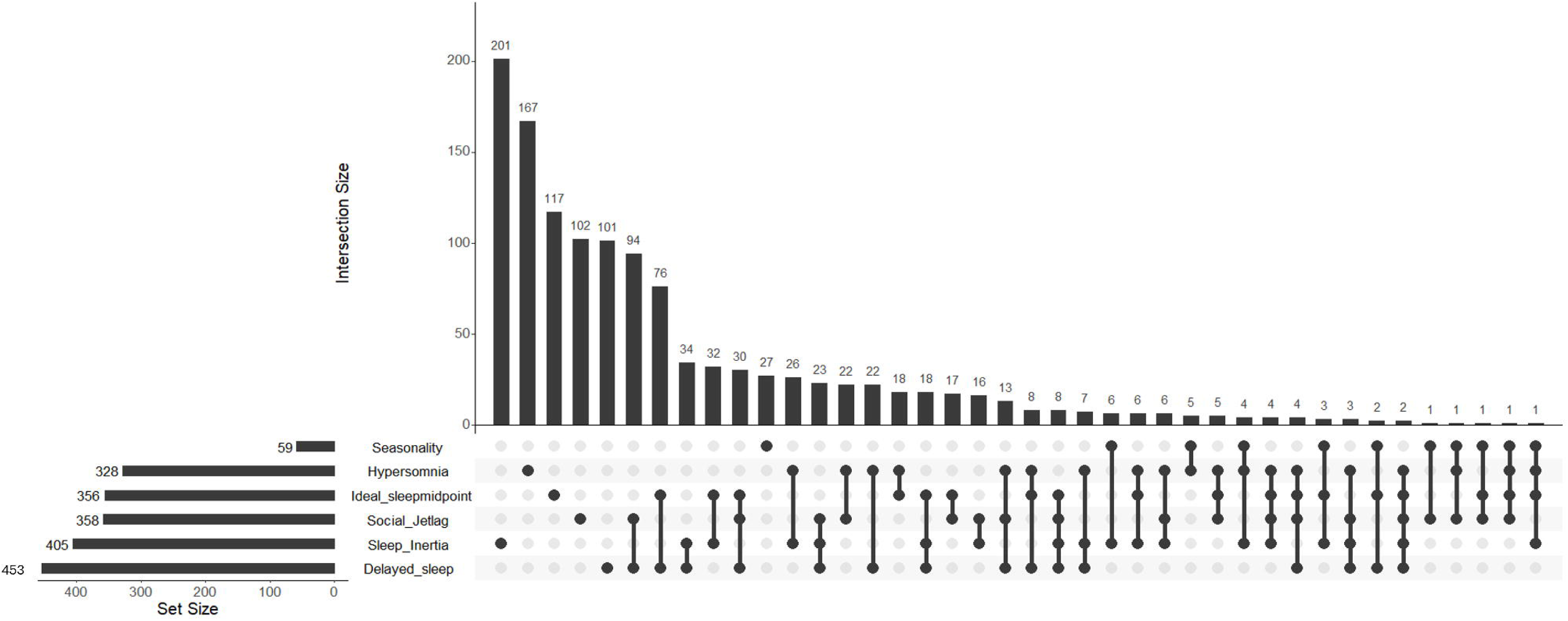
Distributions of circadian-associated phenotypes in participants who endorsed at least one CRD phenotype (n=1,305)

**Table 1.**
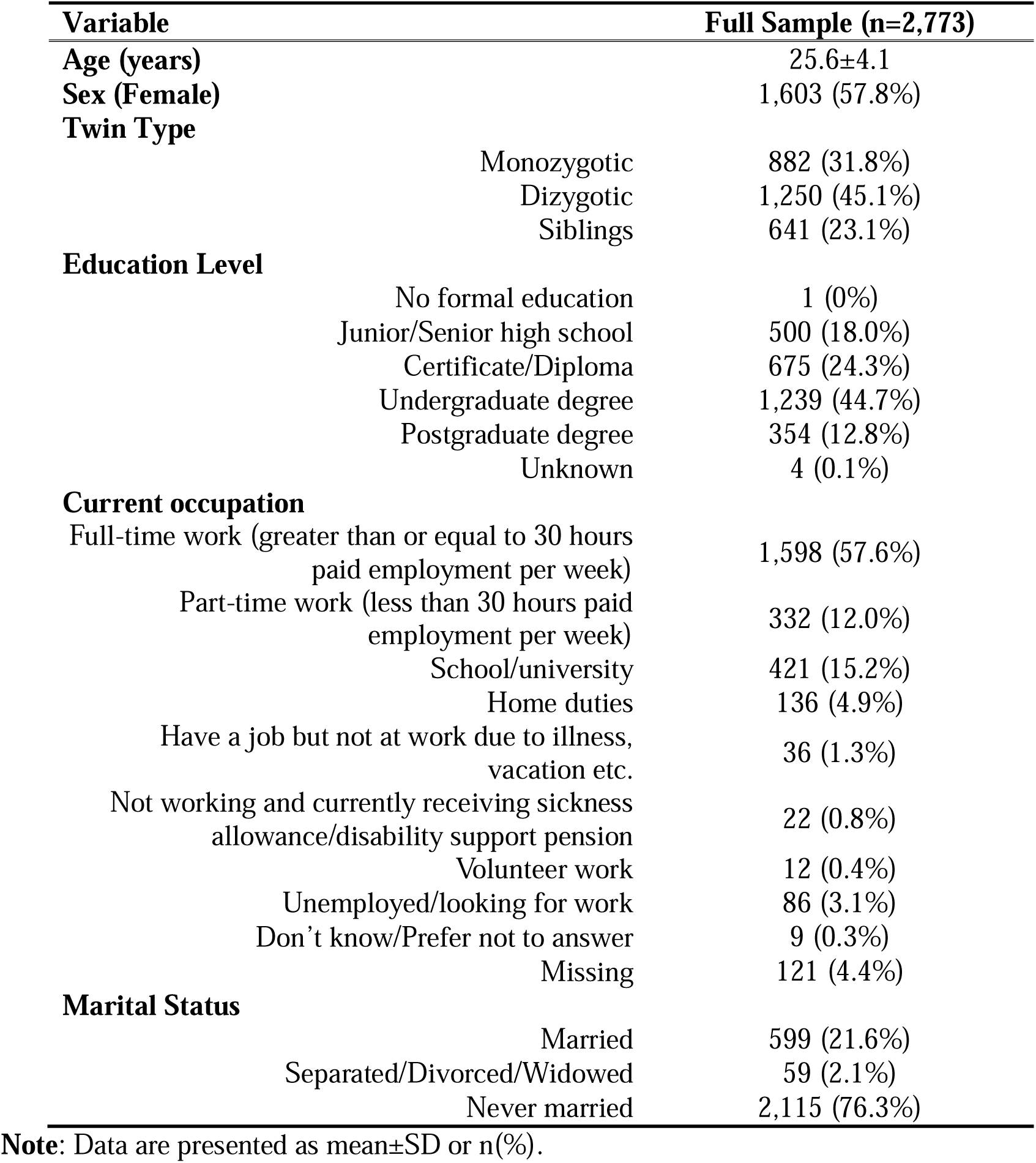
Demographic characteristics of the Brisbane Longitudinal Twin Study sample.

Tetrachoric correlations were generally low, with only timing-related pairs reaching moderate values (delayed sleep-social jetlag, r=0.56; delayed sleep-evening preference, r=0.50; all other pairs, r=-0.09 to 0.27), indicating distinct but related aspects of CRD (Table S4). Seasonality completion was 22.5%, reflecting the data-collection phase described above; participant characteristics by completion status are reported in Table S5.

Demographic and clinical characteristics across CRD-load levels are compared descriptively in Table S6. Higher CRD-load was associated with younger age (OR=0.94 [0.92-0.96], p<0.001) and male sex (OR=1.18 [1.01-1.38], p=0.038), and lower CRD-load with being married (OR=0.60 [0.49-0.74], p<0.001). Education and twin status were not significant predictors (Table S7).

### Clinical and functioning correlates

Higher CRD-load was significantly associated with worse outcomes across all clinical and functional domains tested (Table 2). Each additional CRD phenotype was associated with a 0.96-point increase in K6 scores (β=0.96 [0.72-1.19], p<0.001); a 40% higher count of psychotic-like experiences (IRR=1.40 [1.25-1.58], p<0.001); a 7% higher count of hypomanic-like experiences (IRR=1.07 [1.02-1.12], p=0.003); 31% higher somatic symptoms (IRR=1.31 [1.24-1.39], p<0.001); and 28% higher psychological symptoms (IRR=1.28 [1.19-1.37], p<0.001). To examine whether this association was driven by overlap with sleep symptoms, we repeated the analysis after excluding the two sleep-related SPHERE somatic items (“Needing to sleep longer” and “Poor sleep”). The association was unchanged (mean score: 0.79±1.48, IRR=1.31[1.20-1.42], p<0.001). Each additional phenotype was also associated with 55% more days in bed (IRR=1.55 [1.35-1.77], p<0.001), 43% more days out of role (IRR=1.43 [1.27-1.61], p<0.001), and 42% higher odds of having migraine (OR=1.42 [1.18-1.71], p<0.001).

**Table 2.** Clinical correlates of CRD-load (range=0-5).

| **Outcome** | **n** | **Mean±SD /**<br>**n (%)** | **Median [IQR]** | **Adjusted Estimate**<br>**[95% CI]** | **p** |
| --- | --- | --- | --- | --- | --- |
| Psychological Distress (K6)<sup>a</sup> | 1692 | 9.74±4.15 | 8 [7-11] | β=0.96 [0.72, 1.19] | <0.001* |
| Psychotic-like Experiences (CAPE)<sup>b</sup> | 2651 | 0.19±0.65 | 0 [0-0] | IRR=1.40 [1.25, 1.58] | <0.001* |
| Hypomanic-like Experiences (ASRM)<sup>b</sup> | 2651 | 1.60±1.87 | 0 [0-0] | IRR=1.07 [1.02, 1.12] | 0.003* |
| Somatic Symptoms (SPHERE)<sup>b</sup> | 2651 | 1.51±2.36 | 0 [0-2] | IRR=1.31 [1.24, 1.39] | <0.001* |
| Psychological Symptoms (SPHERE)<sup>b</sup> | 2651 | 1.02±2.06 | 0 [0-1] | IRR=1.28 [1.19, 1.37] | <0.001* |
| Days Out of Role (count)<sup>b</sup> | 1699 | 1.23±3.28 | 0 [0-0] | IRR=1.43 [1.27, 1.61] | <0.001* |
| Days in Bed (count)<sup>b</sup> | 1694 | 0.40±1.25 | 0 [0-0] | IRR=1.55 [1.35, 1.77] | <0.001* |
| Having a Migraine<sup>c</sup> | 2651 | 105 (4.0%) | — | OR=1.42 [1.18, 1.71] | <0.001* |

**Note:** Models adjusted for age, sex, and twin status with standard errors clustered on family ID. <sup>a</sup> Using linear regression; <sup>b</sup> Using negative binomial for count-based variables due to skewed/zero-inflated counts; <sup>c</sup> Using logistic regression for binary variables. \*Bonferroni-corrected significance ( $\alpha=0.006$ ). β=unstandardized coefficient; IRR=Incidence Rate Ratio; OR=Odds Ratio; CI = Confidence Interval.

### Heritability of CRD phenotypes

Four CRD phenotypes showed significant additive genetic influence (Table 3). Among these, delayed sleep was the most heritable in both sexes (h^2^=0.48 in females, 0.44 in males), followed by evening preference (0.36/0.41 female/male), hypersomnia (0.32/0.19), and social jetlag (0.22, no sex differences). The supportive weekday sleep-midpoint estimate (used to derive social jetlag) was higher in females (h^2^=0.57). In contrast, seasonality showed no detectable additive genetic influence (best-fitting CE model; c^2^=0.28), and the sleep inertia showed no evidence of heritability or familial aggregation (e^2^=1.00). For sleep inertia, the MZ and DZ twin correlations differed by less than 0.01. Overall, sleep-timing phenotypes showed moderate heritability, whereas seasonality reflected predominantly shared environmental influences and sleep inertia was largely explained by individual-specific environmental factors. Full model-fitting comparisons are reported in SD13-19.

**Table 3.** Heritability estimates for the continuous phenotypes underlying circadian rhythm disturbance (CRD) phenotypes.

| CRD phenotype | Continuous measure modelled | Best model | A [95% CI] |  | C [95%CI] | E (Female/ Male) |
| --- | --- | --- | --- | --- | --- | --- |
|  |  |  | Female | Male |  |  |
| Hypersomnia | Weekend sleep duration | AEq | 0.32 [0.20-0.43] | 0.19 [0.06-0.33] | — | 0.68 / 0.81 |
| Delayed sleep | Weekend sleep midpoint | AEq | 0.48 [0.37-0.57] | 0.44 [0.31-0.56] | — | 0.52 / 0.56 |
| Evening preference | Sleep midpoint on ideal days | AEq | 0.36 [0.25-0.46] | 0.41 [0.28-0.52] | — | 0.64 / 0.59 |
| Social jetlag | Weekend-weekday sleep midpoint | AE | 0.22 [0.14-0.30] |  | — | 0.78 |
| Seasonality | Seasonality (GSS) | CE | — |  | 0.28 [0.13-0.42] | 0.72 |
| Sleep inertia | Sleep inertia | E | 0 |  | 0 | 1 |
| NA | Weekday sleep midpoint<sup>#</sup> | AEq | 0.57 [0.47-0.65] | 0.41 [0.27-0.53] | — | 0.43 / 0.59 |

**Note:** <sup>#</sup> Weekday sleep midpoint is not itself a CRD phenotype; it is reported as a supportive timing estimate and enters the social jetlag calculation (weekend – weekday midpoint). A = additive genetic; C = shared environmental; E = unique environmental (including measurement error). Best-fitting models: AE = additive genetic + unique environmental; AEq = AE with quantitative sex differences in A and E parameters; CE = shared + unique environmental (no additive genetic component); E = unique environmental only. Where the best-fitting model retained sex differences, female and male estimates are presented separately; otherwise, the same estimate applies to both sexes. 95% confidence intervals are shown in parentheses. Dash (—) indicates parameters not retained in the best-fitting model.

### Genetic associations

Genetic analyses included 2,301 participants (Figure 2). Full estimates and covariate effects are reported in SD1-2. Among mental health PRSs, a higher CRD-load was significantly associated with higher PRSs for bipolar disorder (OR=1.14 [1.05-1.24], p=0.002) and major depression (OR=1.14 [1.05-1.23], p=0.003) but not ADHD (OR=1.02 [0.93-1.12], p=0.703), schizophrenia (OR=1.04 [0.96-1.13], p=0.350), autism (OR=1.03 [0.95-1.11], p=0.539), neuroticism (OR=1.01 [0.93-1.10], p=0.816), or anxiety (OR=0.99 [0.91-1.08], p=0.876).

**Figure 2.**
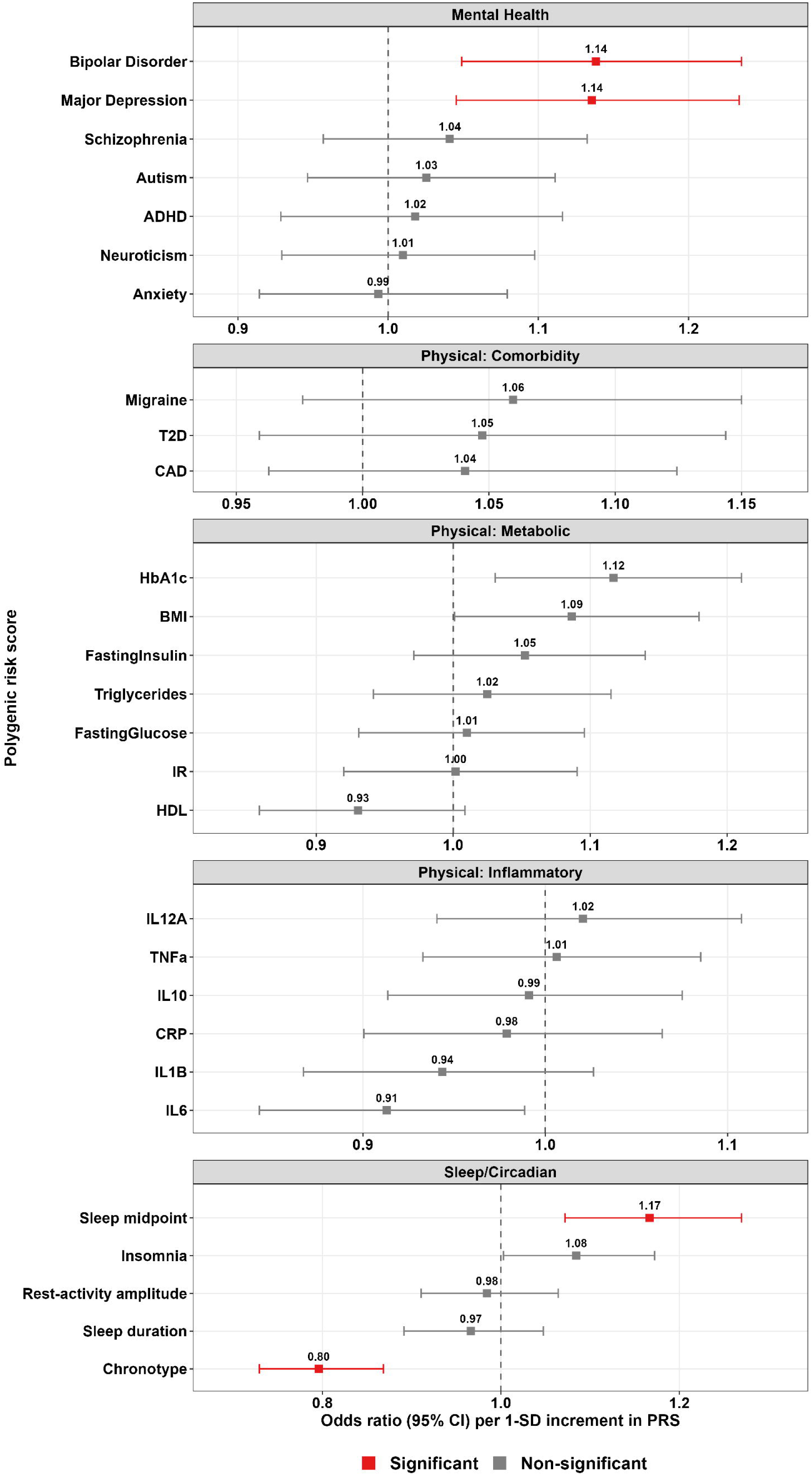
Polygenic risk score associations with CRD-load (range=0-5) across mental health, physical health, and sleep/circadian traits in the full sample (n=2,301). **Note:** Results shown are from separate ordinal logistic regression models for each PRS predicting the number of CRD phenotypes met (range: 0-5), adjusted for age, sex, twin status, and the first four genetically-inferred ancestry PCs, with cluster-robust (sandwich) standard errors clustered on family ID. Bars indicate 95% CI. Red indicates Bonferroni-corrected significance within each domain (mental health: α=0.007; physical health: α=0.003; sleep/circadian: α=0.010); grey indicates non-significant associations. **Abbreviations:** PRS = Polygenic Risk Score; OR = Odds Ratio; CI = Confidence Interval; ADHD = Attention Deficit Hyperactivity Disorder; HbA1c = Hemoglobin A1c; TNF-α = Tumor Necrosis Factor Alpha; IL = Interleukin; BMI = Body Mass Index; T2D = Type 2 Diabetes; CRP = C-Reactive Protein; CAD = Coronary Artery Disease; IR = Insulin Resistance; HDL = High-Density Lipoprotein Cholesterol

Among the 16 physical health related PRS (comorbidities, metabolic-inflammatory, and immune-related traits), no associations survived Bonferroni correction, although glycated hemoglobin (HbA1c, OR=1.12 [1.03-1.21], p=0.007), BMI (OR=1.09 [1.00-1.18], p=0.048), and IL-6 (OR=0.91 [0.84-0.99], p=0.025) had nominal associations.

Among sleep/circadian PRS, a higher CRD-load was associated with lower chronotype PRS, indicating evening chronotype (OR=0.80 [0.73-0.87], p<0.001) and later sleep midpoint (i.e., delayed sleep) PRS (OR=1.17 [1.07-1.27], p<0.001). Insomnia PRS showed a nominal association (OR=1.08 [1.00-1.17], p=0.042).

### Clinical subsample analyses

In the full sample (n=2,651), a greater CRD-load was associated with meeting CIDI criteria for a full-threshold mental disorder (OR=1.28 [1.18-1.39], p<0.001). Among participants meeting CIDI criteria for mental disorders (n=961 of 999 with valid CRD-load data; Table S8&9 and Figure S1), clinical and functional correlates of CRD-load (Table S10) were similar to the full sample. However, age-of-onset was not associated with CRD-load (β=0.08, p=0.691; Table S10). Genetic associations were generally stronger in the clinical subsample (n=808 with genotype data; SD3-4): a higher CRD-load was significantly associated with higher PRS for major depression (OR=1.22), evening chronotype (OR=0.77), and delayed sleep-midpoint (OR=1.37), after Bonferroni correction; the bipolar disorder PRS association was of comparable magnitude (OR=1.20) but did not survive Bonferroni correction in this smaller sample (Figure S2).

### Sensitivity analyses

Among complete responders to all six CRD phenotypes (n=625), no PRS association survived Bonferroni correction; sleep midpoint PRS remained the most strongly associated with CRD-load (OR=1.28 [1.04-1.58], p=0.019), while mental health and chronotype associations were attenuated. Restricting the CRD-load to the four heritable phenotypes (hypersomnia, social jetlag, delayed sleep, evening preference; n=2,301) preserved the significant sleep/circadian associations (evening chronotype OR=0.81 [0.74-0.89], p<0.001; later sleep midpoint OR=1.16 [1.06-1.27], p=0.001) but attenuated the mood disorder PRS associations, with bipolar disorder retained at nominal significance only (OR=1.10 [1.01-1.20], p=0.025) and major depression no longer significant (OR=1.08 [0.99-1.18], p=0.079). In sex-stratified analyses, later chronotype PRS was associated with CRD-load in both sexes; bipolar disorder and sleep midpoint PRS only in females, and insomnia PRS only in males (see Supplementary Materials). Redefining phenotypes using age-specific thresholds shifted prevalences: hypersomnia 16.7% to 12.4%; social jetlag 15.3% to 13.5%; delayed sleep 14.6% to 17.1%; evening preference 12.7% to 13.4%, such that delayed sleep became the most prevalent phenotype and hypersomnia the second least (Table S11). Clinical, functional, and genetic associations were nonetheless unchanged in pattern and magnitude (Tables S11-S12; Figures S6-S7).

## DISCUSSION

In a large community-based twin cohort of young adults with and without mental disorders, greater CRD-load showed a consistent dose-response association with mental health symptoms, functional impairment, and genetic liability for mood disorders and sleep-timing traits. Nearly half of participants met at least one CRD phenotype, broadly consistent with the normative developmental delay in the sleep-wake cycle during adolescence and young adulthood (Logan et al. 2018; Pifer et al. 2024). Together, these findings suggest that a brief set of self-reported features captures clinically meaningful variation in circadian-associated disturbances that is associated with psychopathology and functional impairment.

The association between CRD-load and clinical and functional outcomes was the most robust finding. Each additional CRD phenotype was associated with higher symptom counts, more days-out-of-role and days-in-bed, and higher odds of a full-threshold mental disorder. These findings are consistent with evidence that CRD has broad phenotypic effects spanning cognition, mood and daytime functioning (Logan and McClung 2019; Pifer et al. 2024; Takaesu et al. 2022). Importantly, these associations extended across mood, psychotic-like, somatic and functional domains, supporting the conceptualization of CRD as a transdiagnostic phenotype rather than a marker of any single disorder. The dose-response relationship remained within the clinical subsample and when CRD phenotypes were redefined using age-specific thresholds, which changed individual prevalences but left the clinical and functional associations essentially unchanged, indicating robustness to how phenotypes are defined.

The functional findings warrant particular attention. Functioning during young adulthood shapes educational attainment, employment and social participation, and difficulties in this period predict poorer long-term trajectories (Iorfino et al. 2018). Impairment is already common at first presentation of mood disorders, affecting at least 30% across objective, self-rated and observer-rated domains (Scott et al. 2014). Our findings extend this by showing that impairment increases progressively with cumulative CRD burden. Beyond the cross-sectional associations reported here, CRD has also been shown to predict subsequent functional decline in young people presenting to mental health services (Iorfino et al. 2024). One caveat is that the days-in-bed item refers to time in bed due to illness or injury, and may therefore capture conditions unrelated to CRD (although the same graded pattern was evident for days-out-of-role and symptom counts).

Higher CRD-load was associated with the major depression and bipolar disorder PRSs, but not with schizophrenia, ADHD, autism, neuroticism or anxiety PRSs. This pattern is consistent with models linking circadian disturbance to mood disorder pathophysiology (Hickie et al. 2013; Hickie et al. 2019; McCarthy et al. 2022), although this should be regarded as hypothesis-generating rather than evidence of pathway specificity. We did not formally test whether mood disorder PRS associations exceeded those for other traits, and differences in discovery-GWAS power and in the composition of the BLTS sample, which includes more mood disorder than psychotic disorder cases, may have contributed to the observed pattern. Furthermore, because the cohort comprised young adults, genetic liability to later-onset disorders may not yet be fully expressed.

The same degree of caution applies to the physical health PRSs, none of which survived correction for multiple testing. Metabolic dysfunction is common in the early phases of mood disorders (Shin et al. 2025b; Toyoura et al. 2020) and physical health complications at this age may arise as downstream consequences of circadian disruption and associated behaviors rather than, or in addition to, shared genetic liability, with metabolic-inflammatory contributions becoming more apparent as illness burden, medication exposure and lifestyle factors accumulate (Berk et al. 2023; Correll et al. 2017). Consistent with this, a circadian depression subtype has been associated with metabolic-inflammatory genetic risk in an older cohort with more severe and recurrent depression (Tonini et al. 2026). Differences in ascertainment, severity and sample size preclude direct comparison.

The phenotypic, genetic and twin-model findings together help clarify what CRD-load captures. The relatively low tetrachoric correlations indicate that the six phenotypes represent related but largely distinct circadian characteristics rather than a single underlying construct. Associations with chronotype and sleep-midpoint PRSs support the construct validity of CRD-load, although some convergence is expected because four of the six phenotypes were derived from sleep timing. By contrast, the nominal insomnia association likely reflects shared overlap with general sleep disturbance and arousal-related traits (Jansen et al. 2019; Lane et al. 2019), rather than circadian phase. The absence of an association with the low relative amplitude PRS (which indexes rest-activity rhythm robustness measured via actigraphy (Ferguson et al. 2018)) may reflect both the limits of survey-based assessment and power of this GWAS. Twin modelling was consistent with this pattern: the sleep timing and duration phenotypes were heritable, in line with established estimates for chronotype and sleep timing (Barclay et al. 2010; Gehrman et al. 2019; Jones et al. 2019; Toomey et al. 2015), whereas seasonality was primarily explained by shared environmental influences and sleep inertia by individual-specific environmental factors. Because unique-environmental variance also captures measurement error, the near-zero additive genetic estimate for sleep inertia (a single dichotomized item) may partly reflect limited reliability rather than an absence of genetic contribution. Collectively, these findings suggest that our CRD-load primarily indexes circadian phase-related liability rather than general sleep dysfunction or rhythm robustness.

The sensitivity analyses further inform interpretation of the genetic findings. Restricting CRD-load to the four heritable phenotypes preserved the associations with sleep-timing PRSs but attenuated the associations with mood disorder PRSs. This suggests that the association between CRD-load and mood disorder PRSs is not explained solely by the heritable sleep-timing components of CRD-load and may also reflect information captured by the broader multidimensional construct. However, because this sensitivity analysis removed both seasonality and sleep inertia simultaneously, we cannot determine whether either phenotype, or both, contributed to the attenuation. These exploratory findings therefore require replication and direct modelling of phenotype-specific genetic associations before stronger conclusions can be drawn.

Excluding the two sleep-related SPHERE-12 items left the clinical associations essentially unchanged, reducing the concern about circularity. However, we did not examine whether CRD-load provides predictive information beyond existing symptom measures. Establishing its incremental value for identifying individuals at risk of poorer clinical or functional outcomes will require prospective studies that directly compare CRD-load with established psychiatric assessments. Nevertheless, the robust dose-response relationships observed across symptoms and functional domains suggest that even subclinical accumulation of CRD may have clinical relevance. Given the simplicity of these self-report measures, CRD-load could provide a scalable approach for circadian-associated screening within the emerging field of chronopsychiatry (Smith et al. 2024), where CRDs are increasingly recognized as candidate biomarkers relevant to mental disorders (Crouse et al. 2026).

Several limitations warrant consideration. First, the BLTS survey was not designed to assess CRDs, so the phenotypes were derived from available measures and represent proxy indicators rather than gold-standard circadian assessments (e.g., melatonin, cortisol, core body temperature). Four of six phenotypes were based on overt sleep-wake timing and therefore did not capture internal physiological circadian timing, amplitude or misalignment. Second, the phenotypes were defined using pragmatic distributional thresholds and require validation against objective circadian markers. Third, CRD-load assigns equal weight to each phenotype without formal testing of dimensionality. Future work should examine whether exploratory factor analysis or latent-variable approaches better capture CRD. Fourth, seasonality was assessed only during the earlier phase of data collection and was infrequently endorsed, limiting statistical power. Fifth, sex differences in the twin models and sex-stratified PRS associations were not formally tested and should not be interpreted as evidence of sex moderation. Sixth, the cross-sectional design limits causal inference about whether CRDs precede or follow mood disorder onset. Seventh, the predominantly European-ancestry sample and European-ancestry discovery GWAS limit generalizability and predictive accuracy in other ancestral groups. Eighth, although the prevalence of risk factors, CIDI diagnoses, and comorbidities in the BLTS is broadly comparable to other adolescent and young adult cohort (Costello et al. 2003), the cohort comprised twins and their non-twin siblings rather than a random population sample. Accordingly, twinning and familial relatedness were accounted for as potential confounders in all analyses.

Priorities for future work are 1) replication of this dimensional approach in independent cohorts; 2) integration of objective circadian measures (e.g., actigraphy, circadian physiology) to validate the survey-based approach and capture additional circadian dimensions such as rhythm amplitude and internal circadian misalignment; 3) longitudinal designs beginning before illness onset to test whether CRD-load predicts onset and course; and 4) identification of the circadian features that best predict treatment response, for which trials in recent-onset samples remain scarce (de Haan et al. 2026).

In summary, this exploratory cross-sectional study demonstrates that cumulative self-reported circadian-associated disturbance is associated with progressively greater clinical symptoms, functional impairment, and genetic liability for mood disorders and sleep-timing traits in young adults. Together, these findings support further evaluation of brief, scalable survey-based measures of circadian-associated disturbance as tools for identifying young people with greater clinical burden and for informing circadian-focused prevention and early intervention strategies.

## Data Availability

All data produced in the present study are available upon reasonable request to the authors

## Acknowledgements

We are indebted to all of the Brisbane Longitudinal Twin Study participants for giving their time to contribute to this study. We thank all the people who helped in the conception, implementation, beta testing, media campaign, and data cleaning. We would like to thank the research participants and employees of 23andMe Research Institute for making this work possible. This study was supported in the form of funding by the National Health and Medical Research Council (NHMRC; Grant/Award Numbers 1031119, 1049911, APP10499110) awarded to NM. This work was further supported by NHMRC EL1 Investigator Grants awarded to JJC and BLM (GNT2008196 and GNT2017176, respectively); a Wellcome Trust Mental Health Award (227089/Z/23/Z); NHMRC L1 and L2 Investigator Grants (APP117291, APP2025674) awarded to SEM; and an NHMRC L3 Investigator Grant (GNT2016346) awarded to IBH. This work was supported in part by a NARSAD Young Investigator Grant from the Brain & Behavior Research Foundation award to JJC.

