## Supplementary Materials for "Cumulative Indicators of Circadian Rhythm Disturbance and its Clinical, Functional and Genetic Correlates in the Brisbane Longitudinal Twin Study"

Supporting material is provided in two files. This document (Supplementary Materials) contains questionnaire items, descriptive tables and figures (Tables S1-S12, Figures S1-S7). Full numerical output is provided separately in the Supplementary Data spreadsheet, whose sheets are labelled SD1-SD19: polygenic risk score (PRS) associations and their covariate effects in the full sample (SD1, SD2), the clinical subsample (SD3, SD4) and complete responders (SD5, SD6); the four-phenotype heritable CRD count (SD7, SD8); sex-stratified analyses (SD9, SD10); age-specific CRD thresholds (SD11, SD12); and twin model-fitting comparisons for each of the seven continuous measures (SD13-SD19).

**Table S1.** Questions and answer options for clinical symptom, and functioning

|  | **Question** | **Answer options** |
| --- | --- | --- |
|  | *Over the past few weeks have you been troubled by:* |  |
| **Anxious-Depressive (SPHERE-12)** | - Feeling nervous or tense | *“never or some of the time”*  *“a good part of the time”*  *“most of the time”* |
|  | - Feeling unhappy and depressed |  |
|  | - Feeling constantly under strain |  |
|  | - Everything getting on top of you |  |
|  | - Losing confidence |  |
|  | - Being unable to overcome difficulties |  |
| **Somatic (SPHERE-12)** | - Muscle pain after activity |  |
|  | - Needing to sleep longer |  |
|  | - Prolonged tiredness after activity |  |
|  | - Poor sleep |  |
|  | - Poor concentration |  |
|  | - Tired muscles after activity |  |
|  | *Have you ever...* |  |
| **Psychotic-like (CAPE)** | - Felt as if the thoughts in your head were not your own? | *“yes”*  *“no”* |
|  | - Heard voices talking to each other when you were alone? |  |
|  | - Heard voices when you were alone? |  |
|  | - Felt that many people around you might hurt or harm you in some way? |  |
|  | - Felt as if many people around you were plotting against you? |  |
|  | - Felt as if the thoughts in your head are being taken away from you? |  |
|  | *Have you ever experienced a definite period where for more than two (2) or three (3) days:* |  |
| **Hypo-manic-like (ASRM)** | - You felt much happier or more cheerful than usual? | *“yes”*  *“no”* |
|  | - You felt much more self-confident than usual? |  |
|  | - You needed much less sleep than usual? |  |
|  | - You talked much more than usual? |  |
|  | - You were much more active than usual? |  |
|  | *In the past 4 weeks, how often did you feel* |  |
| **Distress (K6)** | - Nervous? | *“none of the time”*  *“a little of the time”*  *“some of the time”*  *“most of the time”*  *”all of the time”* |
|  | - Hopeless? |  |
|  | - Restless and fidgety? |  |
|  | - That everything was an effort? |  |
|  | - So sad that nothing could cheer you up? |  |
|  | - Worthless? |  |
|  | *During the last few weeks how many days in total:* |  |
| **Functioning** | - Were you unable to carry out your usual daily activities fully? | *Numeric number* |
|  | - Did you stay in bed all or most of the day because of illness or injury? |  |

**Table S2.** Polygenic risk scores (PRS) examined

| **Trait** | **GWAS citation** |
| --- | --- |
| **Psychiatric/neurological** |  |
| Attention-deficit/hyperactivity disorder (ADHD) | (Demontis et al. 2023) |
| Autism | (Grove et al. 2019) |
| Anxiety | (Strom et al. 2026) |
| Bipolar disorder (BD) | (O'Connell et al. 2025) |
| Major depression | (Adams et al. 2025) |
| Neuroticism | (Schwaba et al. 2025) |
| Schizophrenia | (Trubetskoy et al. 2022) |
| **Metabolic-inflammatory traits** |  |
| Body mass index | (Yengo et al. 2018) |
| Coronary artery disease | (Aragam et al. 2022) |
| Fasting insulin | (Dupuis et al. 2010) |
| Fasting glucose | (Lagou et al. 2021) |
| Glycated hemoglobin (HbA1c) | (Sinnott-Armstrong et al. 2021) |
| HDL-Cholesterol | (Sinnott-Armstrong et al. 2021) |
| Insulin Resistance | (Oliveri et al. 2024) |
| Triglycerides | (Sinnott-Armstrong et al. 2021) |
| Type 2 diabetes | (Suzuki et al. 2024) |
| C-reactive protein | (Said et al. 2022) |
| Interleukin-6 (IL-6) | (Sun et al. 2023) |
| Interleukin-10 (IL-10) | (Sun et al. 2023) |
| Interleukin-12p70 (IL-12) | (Sun et al. 2023) |
| Interleukin-1β (IL-1β) | (Sun et al. 2023) |
| Migraine | (Hautakangas et al. 2022) |
| Tumor necrosis factor-α (TNF-α) | (Sun et al. 2023) |
| **Sleep-circadian traits** |  |
| Sleep midpoint | (Jones et al. 2019) |
| Relative amplitude | (Ferguson et al. 2018) |
| Sleep duration | (Jansen et al. 2019) |
| Insomnia | (Jansen et al. 2019) |
| Chronotype | (Jones et al. 2019) |

**Circadian rhythm disturbance phenotypes**

Circadian rhythm disturbance (CRD) was assessed using available sleep and circadian-related variables from the Brisbane Longitudinal Twin Study (BLTS) dataset. As the original survey was not specifically designed to assess CRD, we utilized existing measures that captured key circadian domains. Six phenotypes were selected representing different aspects of circadian disruption, commonly observed in mood disorders. For continuous variables, a threshold of +1 standard deviation (SD) above the sample mean was applied to identify individuals with elevated CRD in that domain. Table S3 summarizes the operationalization and prevalence of each phenotype.

**1. Seasonality:** Moderate to severe seasonal affective patterns were defined as a Global Seasonality Score (GSS) ≥11 on the Seasonal Pattern Assessment Questionnaire (Rosenthal et al. 1984). GSS data were available for 625 of 2,773 participants (22.5%). Among those with available data, 59 participants (9.4%) met the seasonality criterion.

**2. Hypersomnia:** Hypersomnia was defined as weekend sleep duration greater than +1SD above the sample mean. Among 2,650 participants with available sleep duration data (mean=8.53h, SD=1.33), the corresponding threshold (mean+1SD=9.86h) was rounded to 10 hours for clinical interpretability. A total of 442 participants (16.7%) met this criterion.

**3. Delayed sleep midpoint:** Delayed sleep phase was defined as weekend sleep midpoint later than +1SD above the sample mean. Weekend sleep midpoint reflects natural sleep preferences without work or school constraints. Among 2,650 participants with available data (mean=27.71 h, SD=1.38), the threshold was set at mean+1SD (29.09 h, rounded to 29.25 h, corresponding to 5:15 AM). A total of 387 participants (14.6%) met this criterion.

**4. Social jetlag:** Social jetlag reflects chronic circadian disruption due to misalignment between biological and social sleep timing, operationalized as the difference in sleep midpoint between free days (weekends) and workdays. Among 2,650 participants with available data (mean=1.12h, SD=1.04), the threshold was set at mean+1SD (2.16h, rounded to 2.25h). A total of 405 participants (15.3%) met this criterion.

**5. Evening preference:** No validated chronotype questionnaire was administered in this study. As a proxy for chronotype preference, we used the midpoint of sleep on ideal days, derived from self-reported ideal sleep and wake times. Among 2,651 participants with available data (mean=26.64h, SD=0.96), the threshold was set at mean+1SD (27.60 h which was rounded to 27.75h, corresponding to 3:45 AM). A total of 337 participants (12.7%) met this criterion.

**6. Sleep inertia:** Sleep inertia was assessed through a single question: *“Over the past week, how did you feel when you woke up?”.* Response options were: Wide awake (n=103, 3.9%), Awake (n=701, 26.4%), Still a bit sleepy (n=1,442, 54.4%), and Very sleepy (n=405, 15.3%). Participants who selected “Very sleepy” were classified as experiencing sleep inertia. Of 2,651 participants who responded, 405 (15.3%) met this criterion.

**Table S3.** Summary of CRD phenotypes

| **Criterion** | **Definition** | **n with data** | **Threshold** | **n met (%)** |
| --- | --- | --- | --- | --- |
| Seasonality | GSS ≥11 | 625 | ≥11 | 59 (9.4%)* |
| Hypersomnia | Weekend sleep duration ≥+1SD | 2,650 | ≥10 hours | 442 (16.7%) |
| Delayed sleep | Weekend sleep midpoint ≥+1SD | 2,650 | ≥5:15am | 387 (14.6%) |
| Social jetlag | Weekend-weekday midpoint difference ≥+1SD | 2,650 | ≥2.25 hours | 405 (15.3%) |
| Evening preference | Ideal sleep midpoint ≥+1SD | 2,651 | ≥3:45am | 337 (12.7%) |
| Sleep inertia | “Very sleepy” upon waking | 2,651 | Categorical | 405 (15.3%) |

**Note:** GSS = Global Seasonality Score. * Percentages are of participants with available data for that phenotype. For continuous variables, thresholds were set at +1SD above the sample mean, unless otherwise noted.

Tetrachoric correlations were examined to assess the internal structure of the CRD-load measure (Table S4). In the full sample, most inter-phenotype correlations were low (range: -0.09 to 0.27), with moderate correlations between timing-related phenotypes: delayed sleep-social jetlag (r=0.56) and delayed sleep-evening preference (r=0.50). This pattern was consistent in the clinical subsample (r=0.52 and r=0.44, respectively). The moderate correlations among timing phenotypes reflect shared circadian phase-delay biology expressed through different measures (intrinsic preference, behavioral timing, weekday-weekend mismatch), while the generally low inter-phenotype correlations confirm that individual phenotypes contribute unique variance to the composite CRD-load score. Seasonality was largely independent from timing phenotypes in both samples. Wider confidence intervals for seasonality pairs reflect the smaller sample with available GSS data.

**Table S4.** Tetrachoric correlation matrix among CRD phenotypes: full sample vs clinical subsample

| **Phenotype Pair** | **r [95% CI] (Full)** | **r [95% CI] (Clinical)** |
| --- | --- | --- |
| Delayed Sleep - Social Jetlag | **0.56 [0.49, 0.62]** | **0.52 [0.40, 0.62]** |
| Delayed Sleep - Evening Preference | **0.50 [0.43, 0.56]** | **0.44 [0.31, 0.55]** |
| Seasonality - Hypersomnia | **0.27 [0.06, 0.43]** | **0.42 [0.15, 0.65]** |
| Seasonality - Sleep Inertia | **0.27 [0.07, 0.43]** | **0.31 [0.01, 0.56]** |
| Evening Preference - Social Jetlag | **0.15 [0.06, 0.24]** | 0.12 [-0.04, 0.26] |
| Evening Preference - Sleep Inertia | **0.15 [0.06, 0.23]** | **0.15 [0.02, 0.28]** |
| Delayed Sleep - Sleep Inertia | **0.12 [0.04, 0.21]** | **0.16 [0.03, 0.30]** |
| Hypersomnia - Social Jetlag | **0.12 [0.03, 0.20]** | 0.10 [-0.05, 0.23] |
| Evening Preference - Hypersomnia | **0.09 [0.00, 0.17]** | 0.09 [-0.07, 0.22] |
| Hypersomnia - Sleep Inertia | 0.07 [-0.02, 0.15] | 0.09 [-0.03, 0.22] |
| Social Jetlag - Sleep Inertia | 0.06 [-0.03, 0.14] | 0.00 [-0.13, 0.13] |
| Seasonality - Evening Preference | 0.04 [-0.23, 0.25] | -0.22 [-0.51, 0.08] |
| Hypersomnia - Delayed Sleep | 0.02 [-0.08, 0.10] | 0.08 [-0.05, 0.22] |
| Seasonality - Delayed Sleep | -0.05 [-0.33, 0.16] | -0.23 [-0.53, 0.08] |
| Seasonality - Social Jetlag | -0.09 [-0.33, 0.10] | -0.12 [-0.49, 0.19] |

**Note**: Values are tetrachoric correlations with bootstrapped 95% CIs (1,000 iterations). Pairs ordered by full sample correlation (descending).

**Comparison of participants with and without seasonality assessment**

Seasonality was assessed only during the computer-assisted telephone interview phase (2009-2011) and was not carried over when data capture moved to the online survey phase (2012-2016). Participants with and without GSS data therefore correspond to the two data-collection phases rather than to differential completion of an optional module.

Of the 2,773 participants, 625 (22.5%) had seasonality data (GSS; Table S5). Those with GSS data were slightly older (26.9 vs 25.2 years, p<0.001) and reported higher levels of psychological distress (K6: 10.33 vs 9.40, p<0.001), somatic symptoms (2.35 vs 1.19, p<0.001), and psychological symptoms (1.68 vs 0.77, p<0.001). They also reported more hypomanic-like experiences (2.14 vs 1.36, p<0.001) and psychotic-like experiences (0.36 vs 0.12, p<0.001). Additionally, participants with GSS data were more likely to report functional impairment, including days out of role (33.0% vs 21.7%, p<0.001) and days spent in bed due to illness (22.6% vs 14.6%, p<0.001). Sex distribution did not differ between groups (58.7% vs 57.5% female, p=0.632).

Participants assessed during the telephone phase were older, more symptomatic and more functionally impaired than those assessed online. Seasonality-specific findings should be interpreted with this in mind, as the subsample with GSS data is more symptomatic than the cohort as a whole.

**Table S5.** Characteristics of participants by Global Seasonality Score (GSS) availability

|  | Has GSS Score | Missing GSS Score | test | p |
| --- | --- | --- | --- | --- |
| n | 625 | 2148 |  |  |
| Age (mean±SD) | 26.92±2.91 | 25.22±4.34 | t = 11.41,  df = 1508.1 | <0.001 |
| Female (n (%)) | 367 (58.7) | 1236 (57.5) | χ^2^ = 0.23 | 0.632 |
| K6 (mean±SD) | 10.33±4.05 | 9.40±4.17 | t = 4.54,  df = 1336.5 | <0.001 |
| Soma 6 (mean±SD) | 2.35±2.46 | 1.19±2.22 | t = 10.61,  df = 938.49 | <0.001 |
| Psych 6 (mean±SD) | 1.68±2.35 | 0.77±1.88 | t = 8.89,  df = 867.95 | <0.001 |
| HMLE (mean±SD) | 2.14±1.87 | 1.36±1.82 | t = 9.28,  df = 991.24 | <0.001 |
| PLE (mean±SD) | 0.36±0.88 | 0.12±0.54 | t = 6.48,  df = 764.39 | <0.001 |
| days out role (n (%)) | 206 (33.0) | 259 (21.7) | χ^2^ = 26.58 | <0.001 |
| days in bed (n (%)) | 141 (22.6) | 173 (14.6) | χ^2^ = 17.67 | <0.001 |

**Note**: Continuous variables are presented as mean±SD and compared using Welch’s t-test. Categorical variables are presented as n (%) and compared using chi-square test. GSS = Global Seasonality Score; K6 = Kessler-6 Psychological Distress Scale; HMLE = Hypomanic-like experiences; PLE = Psychotic-like experiences.

**Table S6.** Descriptive comparison of demographic and clinical characteristics across CRD-load levels (0, 1, 2, 3+)

| **Characteristic** | **Overall** | **0** | **1** | **2** | **3+** | **Significance** |
| --- | --- | --- | --- | --- | --- | --- |
|  | N = 2,651 | N = 1,346 | N = 762 | N = 383 | N = 160 |  |
| **Age, years** | 25.8±4.0 | 26.4±4.1 | 25.4±3.8 | 25.0±4.0 | 24.3±3.7 | 0 vs 1; 0 vs 2; 0 vs 3+ |
| **Sex** |  |  |  |  |  | 0 vs 2 |
| F | 1,534 (57.9%) | 805 (59.8%) | 440 (57.7%) | 204 (53.3%) | 85 (53.1%) |  |
| M | 1,117 (42.1%) | 541 (40.2%) | 322 (42.3%) | 179 (46.7%) | 75 (46.9%) |  |
| **Zygosity** |  |  |  |  |  |  |
| MZ | 851 (32.1%) | 457 (34.0%) | 222 (29.1%) | 116 (30.3%) | 56 (35.0%) |  |
| DZ | 1,178 (44.4%) | 587 (43.6%) | 345 (45.3%) | 183 (47.8%) | 63 (39.4%) |  |
| Siblings | 622 (23.5%) | 302 (22.4%) | 195 (25.6%) | 84 (21.9%) | 41 (25.6%) |  |
| **Education** |  |  |  |  |  | 0 vs 2 |
| No formal education | 1 (0.0%) | 1 (0.1%) | 0 (0.0%) | 0 (0.0%) | 0 (0.0%) |  |
| Junior/Senior high school | 460 (17.4%) | 216 (16.0%) | 139 (18.2%) | 71 (18.5%) | 34 (21.3%) |  |
| Certificate/Diploma | 657 (24.8%) | 349 (25.9%) | 174 (22.8%) | 98 (25.6%) | 36 (22.5%) |  |
| Undergraduate degree | 1,181 (44.5%) | 576 (42.8%) | 354 (46.5%) | 180 (47.0%) | 71 (44.4%) |  |
| Postgraduate degree | 348 (13.1%) | 203 (15.1%) | 93 (12.2%) | 33 (8.6%) | 19 (11.9%) |  |
| Unknown | 4 (0.2%) | 1 (0.1%) | 2 (0.3%) | 1 (0.3%) | 0 (0.0%) |  |
| **Current occupation** |  |  |  |  |  | 0 vs 2 |
| Full-time work | 1,595 (60.2%) | 821 (61.0%) | 457 (60.0%) | 220 (57.4%) | 97 (60.6%) |  |
| Part-time work | 332 (12.5%) | 179 (13.3%) | 87 (11.4%) | 46 (12.0%) | 20 (12.5%) |  |
| Employed, not currently at work | 36 (1.4%) | 18 (1.3%) | 8 (1.0%) | 10 (2.6%) | 0 (0.0%) |  |
| Student | 421 (15.9%) | 188 (14.0%) | 129 (16.9%) | 74 (19.3%) | 30 (18.8%) |  |
| Home duties | 136 (5.1%) | 92 (6.8%) | 36 (4.7%) | 7 (1.8%) | 1 (0.6%) |  |
| Unemployed | 86 (3.2%) | 33 (2.5%) | 26 (3.4%) | 19 (5.0%) | 8 (5.0%) |  |
| Sickness allowance/DSP | 22 (0.8%) | 5 (0.4%) | 11 (1.4%) | 3 (0.8%) | 3 (1.9%) |  |
| Volunteer work | 12 (0.5%) | 5 (0.4%) | 4 (0.5%) | 2 (0.5%) | 1 (0.6%) |  |
| (Missing) | 11 (0.4%) | 5 (0.4%) | 4 (0.5%) | 2 (0.5%) | 0 (0.0%) |  |
| **Marital status** |  |  |  |  |  | 0 vs 1; 0 vs 2; 0 vs 3+ |
| Never married | 2,000 (75.4%) | 926 (68.8%) | 609 (79.9%) | 325 (84.9%) | 140 (87.5%) |  |
| Married | 593 (22.4%) | 383 (28.5%) | 143 (18.8%) | 50 (13.1%) | 17 (10.6%) |  |
| Separated/Divorced/Widowed | 58 (2.2%) | 37 (2.7%) | 10 (1.3%) | 8 (2.1%) | 3 (1.9%) |  |
| **K6 distress** | 9.7±4.1 | 9.0±3.3 | 10.1±4.3 | 11.1±5.1 | 11.7±5.3 | 0 vs 1; 0 vs 2; 0 vs 3+ |
| **Psychotic-like (CAPE)** |  |  |  |  |  | 0 vs 1; 0 vs 2; 0 vs 3+ |
| 0 | 2,353 (88.8%) | 1,239 (92.1%) | 648 (85.0%) | 331 (86.4%) | 135 (84.4%) |  |
| 1 | 191 (7.2%) | 77 (5.7%) | 68 (8.9%) | 31 (8.1%) | 15 (9.4%) |  |
| 2 | 60 (2.3%) | 20 (1.5%) | 28 (3.7%) | 6 (1.6%) | 6 (3.8%) |  |
| 3 | 24 (0.9%) | 7 (0.5%) | 9 (1.2%) | 6 (1.6%) | 2 (1.3%) |  |
| 4 | 10 (0.4%) | 0 (0.0%) | 5 (0.7%) | 4 (1.0%) | 1 (0.6%) |  |
| 5 | 5 (0.2%) | 0 (0.0%) | 1 (0.1%) | 3 (0.8%) | 1 (0.6%) |  |
| 6 | 8 (0.3%) | 3 (0.2%) | 3 (0.4%) | 2 (0.5%) | 0 (0.0%) |  |
| **Hypomanic-like (ASRM)** |  |  |  |  |  | 0 vs 1; 0 vs 2; 0 vs 3+ |
| 0 | 1,278 (48.2%) | 694 (51.6%) | 349 (45.8%) | 172 (44.9%) | 63 (39.4%) |  |
| 1 | 259 (9.8%) | 141 (10.5%) | 63 (8.3%) | 34 (8.9%) | 21 (13.1%) |  |
| 2 | 276 (10.4%) | 128 (9.5%) | 82 (10.8%) | 44 (11.5%) | 22 (13.8%) |  |
| 3 | 263 (9.9%) | 127 (9.4%) | 88 (11.5%) | 31 (8.1%) | 17 (10.6%) |  |
| 4 | 227 (8.6%) | 94 (7.0%) | 75 (9.8%) | 44 (11.5%) | 14 (8.8%) |  |
| 5 | 348 (13.1%) | 162 (12.0%) | 105 (13.8%) | 58 (15.1%) | 23 (14.4%) |  |
| **Somatic (SPHERE)** | 1.5±2.4 | 1.1±1.9 | 1.7±2.5 | 2.1±2.9 | 2.3±2.9 | 0 vs 1; 0 vs 2; 0 vs 3+ |
| **Psychological (SPHERE)** | 1.0±2.1 | 0.8±1.7 | 1.1±2.2 | 1.4±2.5 | 1.5±2.4 | 0 vs 1; 0 vs 2; 0 vs 3+ |
| **Days out of role** | 1.2±3.3 | 0.9±2.8 | 1.3±3.1 | 1.8±3.7 | 2.5±5.3 | 0 vs 1; 0 vs 2; 0 vs 3+ |
| **Days in bed** | 0.4±1.2 | 0.3±1.0 | 0.4±1.1 | 0.7±1.8 | 0.7±2.1 | 0 vs 1; 0 vs 2; 0 vs 3+ |
| **Migraine (ICHD-3)** |  |  |  |  |  | 0 vs 3+ |
| No | 2,546 (96.0%) | 1,302 (96.7%) | 732 (96.1%) | 364 (95.0%) | 148 (92.5%) |  |
| Yes | 105 (4.0%) | 44 (3.3%) | 30 (3.9%) | 19 (5.0%) | 12 (7.5%) |  |

**Note:** Data are presented as mean±SD or n(%); Groups reflect the number of CRD phenotypes met among the 2,651 participants with valid responses to at least two of the six CRD items; 122 participants with insufficient valid responses were excluded. CRD-load was capped at 3+ (levels 3-5 combined; n=160) owing to small numbers at the highest levels (n=19 at 4, n=4 at 5). The final column lists which pairwise comparisons against level 0 (no CRD) reached p<0.05, using independent-samples t-tests for continuous variables (age, K6, SPHERE somatic and psychological, days out of role, days in bed), Mann-Whitney U tests for ordinal variables (CAPE, ASRM, education), and chi-square tests for categorical variables (sex, zygosity, marital status, migraine). These comparisons are descriptive and characterize the sample across CRD-load levels only; CRD-load is modelled as a continuous variable in all primary analyses and is not treated as a categorical grouping. Comparisons are unadjusted for covariates, do not account for the non-independence of twins and siblings, and are not corrected for multiple comparisons; p-values are therefore approximate and not confirmatory.

**Table S7.** Associations between demographic characteristics and CRD-load.

| **Predictor** | **OR [95% CI]** | **p-value** |
| --- | --- | --- |
| Age (per year) | 0.94 [0.92, 0.96] | <0.001 |
| Sex (Ref: Female) | 1.18 [1.01, 1.38] | 0.038 |
| Zygosity (Ref: Dizygotic) |  |  |
| Monozygotic | 0.94 [0.78, 1.13] | 0.519 |
| Sibling | 1.16 [0.97, 1.38] | 0.109 |
| Education (per level, 1-5) | 0.95 [0.87, 1.02] | 0.174 |
| Marital status (Ref: Never married) |  |  |
| Married | 0.60 [0.49, 0.74] | <0.001 |
| Separated/Divorced/Widowed | 0.68 [0.37, 1.22] | 0.194 |

**Note.** Estimates are from a single ordinal logistic regression model with CRD-load (number of CRD phenotypes met, 0-5) as the outcome and all listed predictors entered simultaneously, restricted to the 2,651 participants with valid responses to at least two of the six CRD phenotypes. 95% confidence intervals were estimated using cluster-robust (sandwich) variance estimators clustered on family ID to account for the non-independence of twins and siblings. Odds ratios above 1 indicate higher CRD-load. Age is modelled per year and education as an ordinal score from 1 (no formal education) to 5 (postgraduate degree); “Unknown” education (n=4) was treated as missing. OR = odds ratio; CI = confidence interval.

**Secondary analysis: clinical subsample**

Secondary analyses were restricted to participants meeting clinical mood, anxiety, or psychotic disorder criteria (clinical subsample), to examine whether associations were amplified in those with manifest illness. Of the 2,773 participants, 999 (36.0%) met criteria for a clinical mood, anxiety, or psychotic disorder based on structured diagnostic assessment. The clinical subsample was predominantly female (65.0%) with a mean age of 25.8±4.1 years and mean age of disorder onset of 17.8±6.4 years (Table S8).

**Table S8.** Demographic characteristics of the clinical subsample

| **Variable** | **Clinical Sample (n=999)** |
| --- | --- |
| **Age** | 25.8±4.1 |
| **Sex (Female)** | 649 (65.0%) |
| **Age of onset** | 17.8±6.4 |
| **Twin Type** |  |
| Monozygotic | 306 (30.6%) |
| Dizygotic | 422 (42.2%) |
| Siblings | 271 (27.1%) |
| **Education Level** |  |
| No formal education | 0 (0%) |
| Junior/Senior high school | 194 (19.4%) |
| Certificate/Diploma | 274 (27.4%) |
| Undergraduate degree | 421 (42.1%) |
| Postgraduate degree | 108 (10.8%) |
| Unknown | 2 (0.2%) |
| **Marital Status** |  |
| Married | 197 (19.7%) |
| Separated/Divorced/Widowed | 25 (2.5%) |
| Never married | 777 (77.8%) |

**Note:** Data are presented as mean±SD or n(%)

We repeated the primary analyses within participants meeting criteria for a DSM-IV mental disorder, applying the same CRD phenotype definitions and thresholds as in the full sample. Those providing valid responses to at least two of the six CRD phenotypes were retained, giving an analytic subsample of 961. The prevalence of individual CRD phenotypes was 22.2% for sleep inertia (n=213), 19.0% for hypersomnia (n=183), 17.1% for delayed sleep midpoint (n=164), 15.5% for social jetlag (n=149), and 12.2% for evening preference (n=117). Seasonality, assessed in a smaller subset, was met by 15.5% (32/206). Across all instances of a phenotype being met, sleep inertia and hypersomnia were the most common and seasonality the least (Figure S1). In clinical subsample, 45.1% (n=433) met no CRD phenotype and 54.9% (n=528) met at least one: one phenotype in 29.6% (n=284), two in 17.7% (n=170), three in 6.7% (n=64), four in 0.8% (n=8), and five in 0.2% (n=2); none met all six. Single-phenotype profiles were the most common among participants with any CRD, and co-occurrence of multiple phenotypes was less frequent (Figure S1). Descriptive characteristics across CRD-load levels in the clinical subsample are shown in Table S9.

**Table S9.** Descriptive comparison of demographic and clinical characteristics across CRD-load levels (0, 1, 2, 3+) within clinical subsample

| **Characteristic** | **Overall** | **0** | **1** | **2** | **3+** | **Significance** |
| --- | --- | --- | --- | --- | --- | --- |
|  | N = 961 | N = 433 | N = 284 | N = 170 | N = 74 |  |
| **Age, years** | 26.0±4.0 | 26.5±4.1 | 25.9±4.0 | 25.4±4.0 | 24.6±3.6 | 0 vs 2; 0 vs 3+ |
| **Sex** |  |  |  |  |  | 0 vs 2 |
| F | 622 (64.7%) | 286 (66.1%) | 182 (64.1%) | 105 (61.8%) | 49 (66.2%) |  |
| M | 339 (35.3%) | 147 (33.9%) | 102 (35.9%) | 65 (38.2%) | 25 (33.8%) |  |
| **Zygosity** |  |  |  |  |  |  |
| MZ | 295 (30.7%) | 139 (32.1%) | 78 (27.5%) | 51 (30.0%) | 27 (36.5%) |  |
| DZ | 405 (42.1%) | 180 (41.6%) | 126 (44.4%) | 74 (43.5%) | 25 (33.8%) |  |
| Siblings | 261 (27.2%) | 114 (26.3%) | 80 (28.2%) | 45 (26.5%) | 22 (29.7%) |  |
| **Education** |  |  |  |  |  | 0 vs 3+ |
| No formal education | 0 (0.0%) | 0 (0.0%) | 0 (0.0%) | 0 (0.0%) | 0 (0.0%) |  |
| Junior/Senior high school | 183 (19.0%) | 72 (16.6%) | 52 (18.3%) | 37 (21.8%) | 22 (29.7%) |  |
| Certificate/Diploma | 268 (27.9%) | 122 (28.2%) | 77 (27.1%) | 50 (29.4%) | 19 (25.7%) |  |
| Undergraduate degree | 402 (41.8%) | 183 (42.3%) | 128 (45.1%) | 67 (39.4%) | 24 (32.4%) |  |
| Postgraduate degree | 106 (11.0%) | 55 (12.7%) | 26 (9.2%) | 16 (9.4%) | 9 (12.2%) |  |
| Unknown | 2 (0.2%) | 1 (0.2%) | 1 (0.4%) | 0 (0.0%) | 0 (0.0%) |  |
| **Marital status** |  |  |  |  |  | 0 vs 2; 0 vs 3+ |
| Never married | 740 (77.0%) | 308 (71.1%) | 220 (77.5%) | 144 (84.7%) | 68 (91.9%) |  |
| Married | 196 (20.4%) | 111 (25.6%) | 58 (20.4%) | 22 (12.9%) | 5 (6.8%) |  |
| Separated/Divorced/Widowed | 25 (2.6%) | 14 (3.2%) | 6 (2.1%) | 4 (2.4%) | 1 (1.4%) |  |
| **K6 distress** | 11.6±5.0 | 10.3±4.1 | 12.1±5.0 | 13.0±5.9 | 14.3±5.6 | 0 vs 1; 0 vs 2; 0 vs 3+ |
| **Psychotic-like (CAPE)** |  |  |  |  |  | 0 vs 1; 0 vs 2 |
| 0 | 768 (79.9%) | 370 (85.5%) | 211 (74.3%) | 130 (76.5%) | 57 (77.0%) |  |
| 1 | 112 (11.7%) | 44 (10.2%) | 37 (13.0%) | 21 (12.4%) | 10 (13.5%) |  |
| 2 | 44 (4.6%) | 10 (2.3%) | 23 (8.1%) | 6 (3.5%) | 5 (6.8%) |  |
| 3 | 18 (1.9%) | 7 (1.6%) | 6 (2.1%) | 4 (2.4%) | 1 (1.4%) |  |
| 4 | 9 (0.9%) | 0 (0.0%) | 5 (1.8%) | 4 (2.4%) | 0 (0.0%) |  |
| 5 | 5 (0.5%) | 0 (0.0%) | 1 (0.4%) | 3 (1.8%) | 1 (1.4%) |  |
| 6 | 5 (0.5%) | 2 (0.5%) | 1 (0.4%) | 2 (1.2%) | 0 (0.0%) |  |
| **Hypomanic-like (ASRM)** |  |  |  |  |  | 0 vs 1 |
| 0 | 371 (38.6%) | 185 (42.7%) | 103 (36.3%) | 62 (36.5%) | 21 (28.4%) |  |
| 1 | 80 (8.3%) | 34 (7.9%) | 19 (6.7%) | 14 (8.2%) | 13 (17.6%) |  |
| 2 | 112 (11.7%) | 52 (12.0%) | 31 (10.9%) | 19 (11.2%) | 10 (13.5%) |  |
| 3 | 111 (11.6%) | 49 (11.3%) | 35 (12.3%) | 19 (11.2%) | 8 (10.8%) |  |
| 4 | 99 (10.3%) | 34 (7.9%) | 35 (12.3%) | 23 (13.5%) | 7 (9.5%) |  |
| 5 | 188 (19.6%) | 79 (18.2%) | 61 (21.5%) | 33 (19.4%) | 15 (20.3%) |  |
| **Somatic (SPHERE)** | 2.1±2.9 | 1.6±2.4 | 2.3±2.9 | 2.8±3.4 | 3.3±3.3 | 0 vs 1; 0 vs 2; 0 vs 3+ |
| **Psychological (SPHERE)** | 1.7±2.7 | 1.3±2.4 | 1.8±2.8 | 2.1±3.1 | 2.6±2.9 | 0 vs 1; 0 vs 2; 0 vs 3+ |
| **Days out of role** | 1.9±3.8 | 1.3±3.2 | 1.7±3.4 | 2.8±4.3 | 3.5±6.1 | 0 vs 2; 0 vs 3+ |
| **Days in bed** | 0.6±1.6 | 0.4±1.3 | 0.6±1.2 | 1.0±2.2 | 1.1±2.9 | 0 vs 2 |
| **Migraine (ICHD-3)** |  |  |  |  |  | 0 vs 2; 0 vs 3+ |
| No | 909 (94.6%) | 417 (96.3%) | 268 (94.4%) | 157 (92.4%) | 67 (90.5%) |  |
| Yes | 52 (5.4%) | 16 (3.7%) | 16 (5.6%) | 13 (7.6%) | 7 (9.5%) |  |

**Note:** Data are presented as mean±SD or n(%); Groups reflect the number of CRD phenotypes met among the 961 participants with valid responses to at least two of the six CRD items; 38 participants with insufficient valid responses were excluded. CRD-load was capped at 3+ (levels 3-5 combined; n=74) owing to small numbers at the highest levels (n=8 at 4, n=2 at 5). The final column lists which pairwise comparisons against level 0 (no CRD) reached p<0.05, using independent-samples t-tests for continuous variables (age, K6, SPHERE somatic and psychological, days out of role, days in bed), Mann-Whitney U tests for ordinal variables (CAPE, ASRM, education), and chi-square tests for categorical variables (sex, zygosity, marital status, migraine). These comparisons are descriptive and characterize the sample across CRD-load levels only; CRD-load is modelled as a continuous variable in all primary analyses and is not treated as a categorical grouping. Comparisons are unadjusted for covariates, do not account for the non-independence of twins and siblings, and are not corrected for multiple comparisons; p-values are therefore approximate and not confirmatory.

**Table S10.** Clinical correlates of CRD-load within clinical subsample (range=0-5)

| **Outcome** | **n** | **Mean±SD /**  **n (%)** | **Median [IQR]** | **Adjusted Estimate**  **[95% CI]** | **p** |
| --- | --- | --- | --- | --- | --- |
| Psychological Distress (K6)^a^ | 629 | 11.62±5.00 | 10 [8-14] | β=1.24 [0.83, 1.65] | <0.001* |
| Age of onset ^a^ | 898 | 17.76±6.40 | 18 [14-22] | β=0.08 [-0.30, 0.45] | 0.691 |
| Psychotic-like Experiences (CAPE)^b^ | 961 | 0.35±0.88 | 0 [0-0] | IRR=1.28 [1.12, 1.46] | <0.001* |
| Hypomanic-like Experiences (ASRM)^b^ | 961 | 1.97±1.98 | 2 [0-4] | IRR=1.04 [0.99, 1.11] | 0.141 |
| Somatic Symptoms (SPHERE)^b^ | 961 | 2.05±2.85 | 0 [0-3] | IRR=1.29 [1.19, 1.39] | <0.001* |
| Psychological Symptoms (SPHERE)^b^ | 961 | 1.65±2.66 | 0 [0-3] | IRR=1.23 [1.12, 1.35] | <0.001* |
| Days Out of Role (count) ^b^ | 632 | 1.86±3.82 | 0 [0-2] | IRR=1.37 [1.18, 1.59] | <0.001* |
| Days in Bed (count)^b^ | 631 | 0.63±1.65 | 0 [0-1] | IRR=1.43 [1.19, 1.71] | <0.001* |
| Having a Migraine ^c^ | 961 | 105 (10.9%) | — | OR=1.45 [1.15, 1.83] | 0.002* |

**Note**: Models adjusted for age, sex, and twin status with cluster-robust (sandwich) standard errors clustered on family ID. ^a^ Using linear regression; ^b^ Using negative binomial for count-based variables due to skewed/zero-inflated counts; ^c^ Using logistic regression for binary variables. *Bonferroni-corrected significance (α=0.006). β=unstandardized coefficient; IQR = Interquartile Range; IRR=Incidence Rate Ratio; OR=Odds Ratio; CI = Confidence Interval.

**Figure S1.** Co-occurrence of circadian rhythm disturbance (CRD) phenotypes in the clinical subsample (n=961).

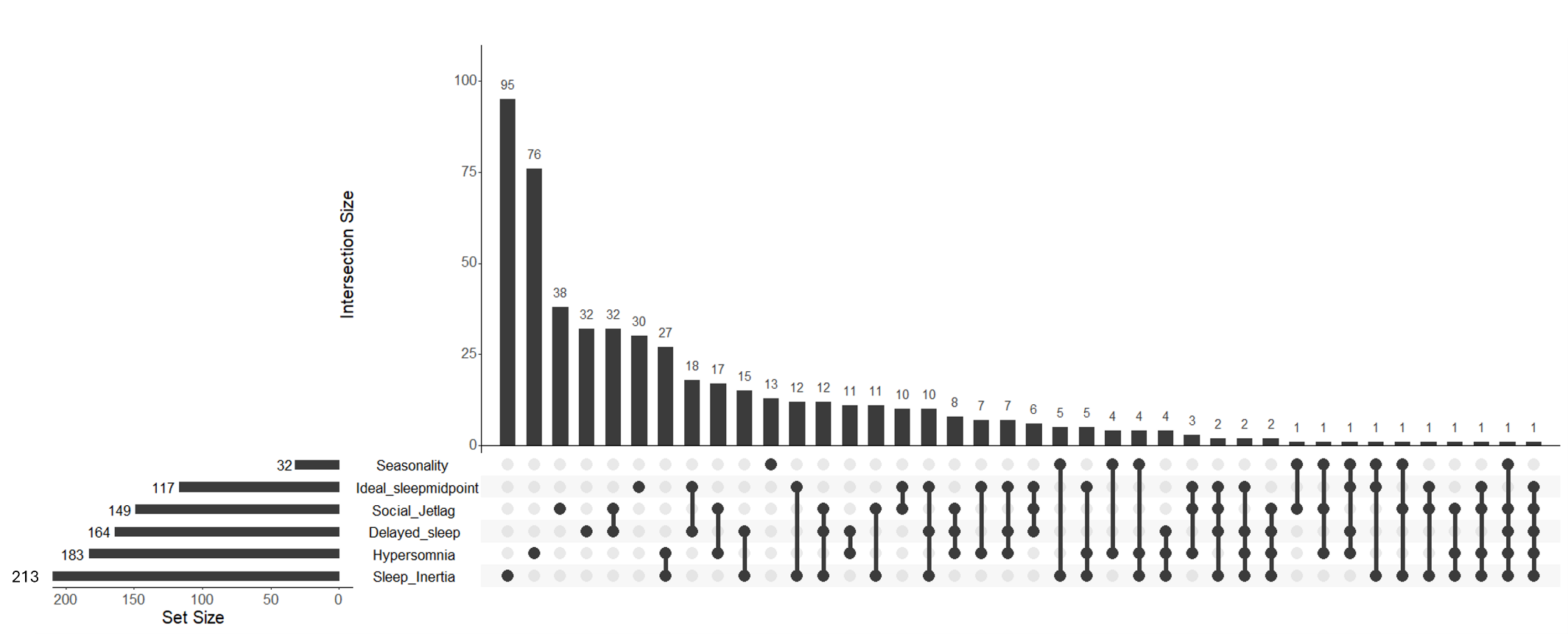

CRD-load was associated with worse clinical outcomes within the clinical subsample (Table S10), with a pattern similar to the primary full-sample analysis. Each additional CRD phenotype was associated with a 1.24-point increase in K6 psychological distress (β=1.24 [0.83-1.65], p<0.001), suggesting a stronger effect than in the full sample (β=0.96). CRD-load was significantly associated with psychotic-like experiences (IRR=1.28, p<0.001), somatic symptoms (IRR=1.29, p<0.001), psychological symptoms (IRR=1.23, p<0.001), days out of role (IRR=1.37, p<0.001), days in bed (IRR=1.43, p<0.001), and migraine (OR=1.45, p=0.002), all surviving Bonferroni correction. Hypomanic-like experiences were not significantly associated (IRR=1.04, p=0.141). Notably, age of onset was not associated with CRD-load (β=0.08, p=0.691), suggesting that CRD-load reflects current symptom severity rather than illness duration.

PRS analyses in this clinical subsample (n=808 with available PRS data; Figure S2) yielded largely consistent results with the primary analysis (Figure 2). Among mental health PRS, higher major depression PRS (OR=1.22 [1.06-1.40], p=0.005) remained significantly associated with greater CRD-load after Bonferroni correction. Bipolar disorder PRS showed an association of comparable magnitude (OR=1.20 [1.04-1.38], p=0.010) that did not survive correction. No other mental health PRS, including anxiety, were significantly associated. Among physical health PRS, no associations survived Bonferroni correction. Sleep/circadian PRS showed stronger effect sizes than in the full sample: lower chronotype PRS (OR=0.77 [0.67-0.89], p<0.001) and higher sleep midpoint PRS (OR=1.37 [1.18-1.58], p<0.001) remained robustly associated with greater CRD-load.

**Figure S2.** Polygenic risk score associations with CRD-load across mental health, physical health, and sleep/circadian domains in the clinical subsample (n=808).

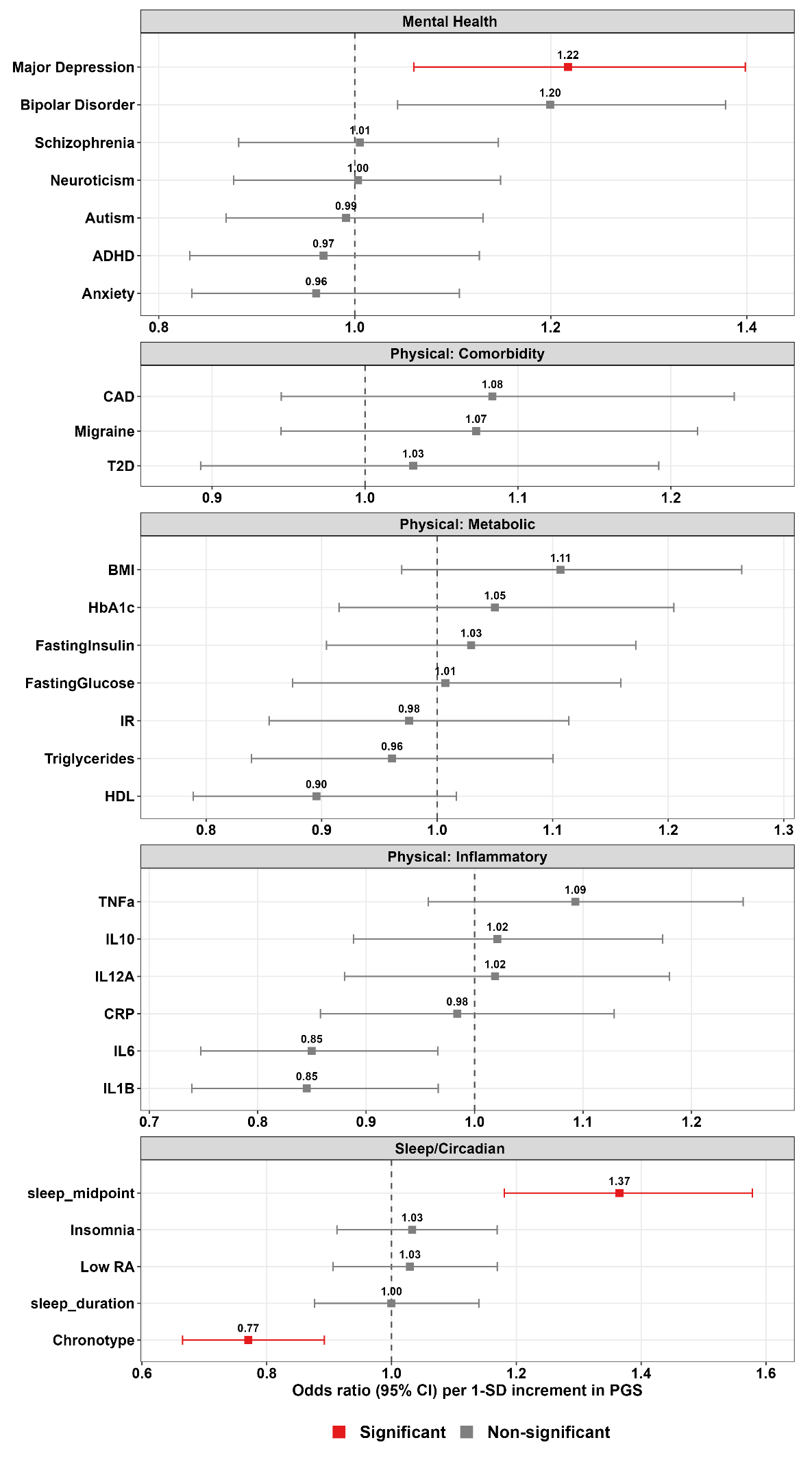

**Note:** Results shown are from separate ordinal logistic regression models for each PRS predicting the number of CRD phenotypes met (range: 0-5), adjusted for age, sex, twin status, and the first four genetically-inferred ancestry PCs, with cluster-robust (sandwich) standard errors clustered on family ID. Bars indicate 95% CI. Red indicates Bonferroni-corrected significance within each domain (mental health: α=0.007; physical health: α=0.003; sleep/circadian: α=0.010); grey indicates non-significant associations.

**Abbreviations:** PRS = Polygenic Risk Score; OR = Odds Ratio; CI = Confidence Interval; ADHD = Attention Deficit Hyperactivity Disorder; HbA1c = Hemoglobin A1c; TNF-α = Tumor Necrosis Factor Alpha; IL = Interleukin; BMI = Body Mass Index; T2D = Type 2 Diabetes; CRP = C-Reactive Protein; CAD = Coronary Artery Disease; IR = Insulin Resistance; HDL = High-Density Lipoprotein Cholesterol; RA = Relative Amplitude

**Sensitivity analyses**

Four sensitivity analyses were conducted to assess the robustness of the primary findings. First, analyses were restricted to complete responders who provided valid responses to all six CRD phenotypes, ensuring that the ordinal outcome captured the full range of CRD-load. Second, the CRD count was restricted to the four phenotypes showing evidence of additive genetic influence (hypersomnia, social jetlag, delayed sleep, evening preference), excluding seasonality and sleep inertia. Third, PRS associations were examined separately in females and males. Fourth, the CRD phenotypes were redefined using thresholds derived separately in participants aged 18-24 years and those aged 25 years and over. The first three sensitivity analyses alter the sample or the composition of the CRD count used in the PRS models, and are each reported as a single figure. The fourth alters the definition of the phenotypes themselves and is therefore carried through prevalence, co-occurrence, clinical and functional outcomes, and PRS associations.

All PRS models in the sensitivity analyses used the same ordinal logistic regression approach as the primary analysis, with the number of CRD phenotypes met as the outcome and adjustment for age, sex (except in the sex-stratified analysis), twin status, and the first four ancestry principal components, with family-clustered (sandwich) standard errors. Bonferroni correction was applied within each domain (psychiatric: α=0.007; physical health: α=0.003; sleep/circadian: α=0.010).

**Sensitivity Results 1: complete responders**

Analyses were restricted to the 625 participants who provided valid responses to all six CRD phenotypes (Figure S3). In this reduced sample, no PRS survived Bonferroni correction. Sleep midpoint PRS remained the strongest signal (OR=1.28 [1.04-1.58], p=0.019) but fell short of the sleep/circadian threshold (α=0.010), and the chronotype association was also attenuated (OR=0.89 [0.74-1.07], p=0.20). Mental health associations from the primary analysis also weakened: major depression (OR=1.24 [1.02-1.51], p=0.034) and neuroticism (OR=1.26 [1.03-1.56], p=0.027) were nominally significant, while bipolar disorder was not (OR=1.08 [0.90-1.31], p=0.41). This attenuation is consistent with the substantially reduced sample size (n=625 vs n=2,301 in the primary analysis; a 73% reduction). Effect directions were largely preserved across domains, indicating that the primary findings were not driven by participants with incomplete CRD reporting; rather, statistical power was reduced.

**Figure S3.** Polygenic risk score associations with CRD-load across mental health, physical health, and sleep/circadian domains among complete responders (n=625).

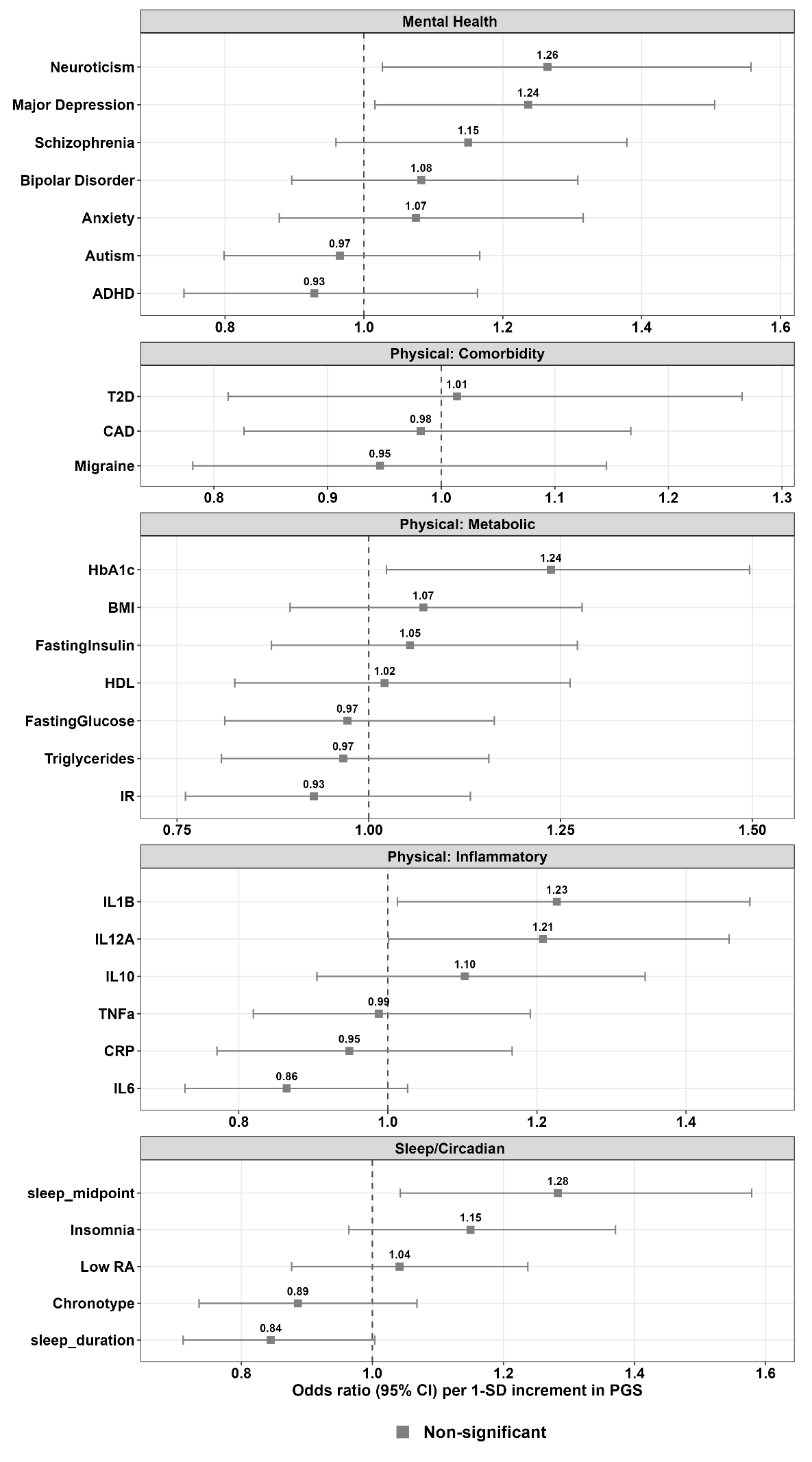

**Note:** Results shown are from separate ordinal logistic regression models for each PRS predicting the number of CRD phenotypes met (range: 0-5), adjusted for age, sex, twin status, and the first four genetically-inferred ancestry PCs, with cluster-robust (sandwich) standard errors clustered on family ID. Bars indicate 95% CI. Red indicates Bonferroni-corrected significance within each domain (mental health: α=0.007; physical health: α=0.003; sleep/circadian: α=0.010); grey indicates non-significant associations.

**Abbreviations:** PRS = Polygenic Risk Score; OR = Odds Ratio; CI = Confidence Interval; ADHD = Attention Deficit Hyperactivity Disorder; HbA1c = Hemoglobin A1c; TNF-α = Tumor Necrosis Factor Alpha; IL = Interleukin; BMI = Body Mass Index; T2D = Type 2 Diabetes; CRP = C-Reactive Protein; CAD = Coronary Artery Disease; IR = Insulin Resistance; HDL = High-Density Lipoprotein Cholesterol; RA = Relative Amplitude

**Sensitivity Results 2: heritable CRD phenotypes** **only**

To test whether genetic associations were robust to the inclusion of phenotypes with negligible additive genetic variance, analyses were repeated using a four-phenotype CRD count restricted to the heritable phenotypes (hypersomnia, delayed sleep, evening preference, and social jetlag), excluding seasonality and sleep inertia (Figure S4). Because all four phenotypes derive from the same sleep-timing items, the analyzable sample (n=2,301) was comparable to the primary analysis, so differences in findings reflect the narrower phenotype rather than reduced power. Sleep/circadian PRS associations were preserved, with chronotype PRS (OR=0.81 [0.74-0.89], p<0.001) and later sleep midpoint PRS (OR=1.16 [1.06-1.27], p<0.001) remaining significant after Bonferroni correction. In contrast, the mood-disorder PRS associations observed in the primary analysis attenuated and no longer survived correction: major depression (OR=1.08 [0.99-1.18], p=0.079) and bipolar disorder (OR=1.10 [1.01-1.20], p=0.025). No physical health PRS reached significance, consistent with the primary analysis. These results indicate that the genetic association between CRD-load and mood-disorder liability reflects the full multi-phenotypes rather than the heritable sleep-timing phenotypes alone.

**Figure S4.** Polygenic risk score associations with the four-phenotype heritable CRD count across mental health, physical health, and sleep/circadian domains (n=2,301).

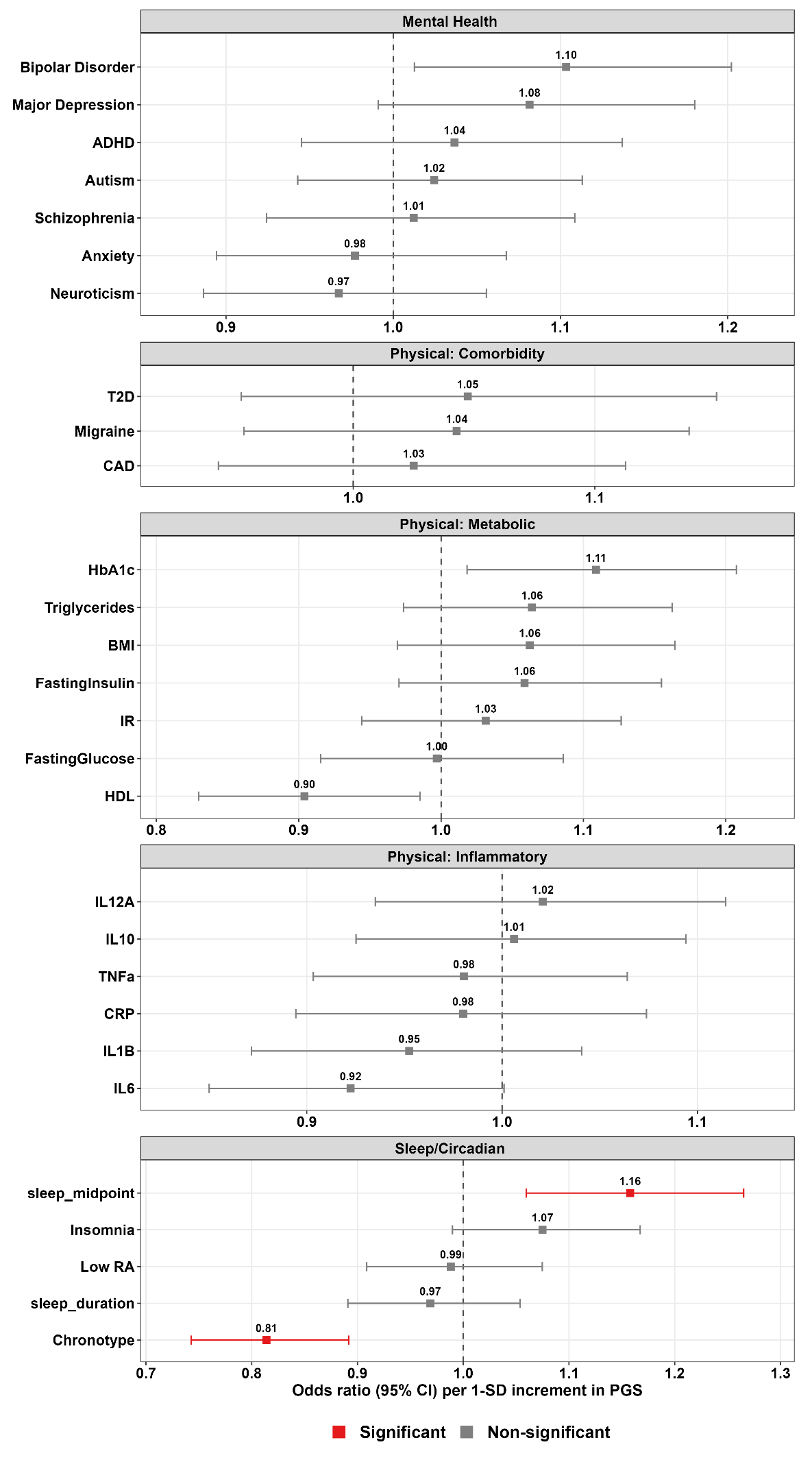

**Note:** Results shown are from separate ordinal logistic regression models for each PRS predicting the number of CRD phenotypes met restricted to the four heritable phenotypes (hypersomnia, social jetlag, delayed sleep, evening preference; range: 0-4), adjusted for age, sex, twin status, and the first four genetically-inferred ancestry PCs, with cluster-robust (sandwich) standard errors clustered on family ID. Bars indicate 95% CI. Red indicates Bonferroni-corrected significance within each domain (mental health: α=0.007; physical health: α=0.003; sleep/circadian: α=0.010); grey indicates non-significant associations.

**Abbreviations**: PRS = Polygenic Risk Score; OR = Odds Ratio; CI = Confidence Interval; ADHD = Attention Deficit Hyperactivity Disorder; HbA1c = Hemoglobin A1c; TNF-α = Tumor Necrosis Factor Alpha; IL = Interleukin; BMI = Body Mass Index; T2D = Type 2 Diabetes; CRP = C-Reactive Protein; CAD = Coronary Artery Disease; IR = Insulin Resistance; HDL = High-Density Lipoprotein Cholesterol; RA = Relative Amplitude

**Sensitivity Results 3: sex-stratified analyses**

PRS associations with CRD-load were examined separately in females and males, with sex omitted from the covariates (Figure S5). Chronotype PRS was associated with CRD-load in both sexes (females OR=0.80 [0.72-0.90]; males OR=0.79 [0.69-0.91]; both p<0.001), indicating a consistent, sex-independent association. Among females, bipolar disorder PRS (OR=1.17 [1.05-1.31], p=0.006) and later sleep midpoint PRS (OR=1.25 [1.11-1.40], p<0.001) were significant after Bonferroni correction, with major depression PRS nominally associated (p=0.018). Among males, insomnia PRS was significant (OR=1.23 [1.09-1.40], p=0.001), whereas the sleep midpoint association was not (OR=1.08 [0.95-1.22], p=0.26). No physical health PRS reached significance in either sex.

**Figure S5**. Sex-stratified polygenic risk score associations with CRD-load across mental health, physical health, and sleep/circadian domains in females and males.

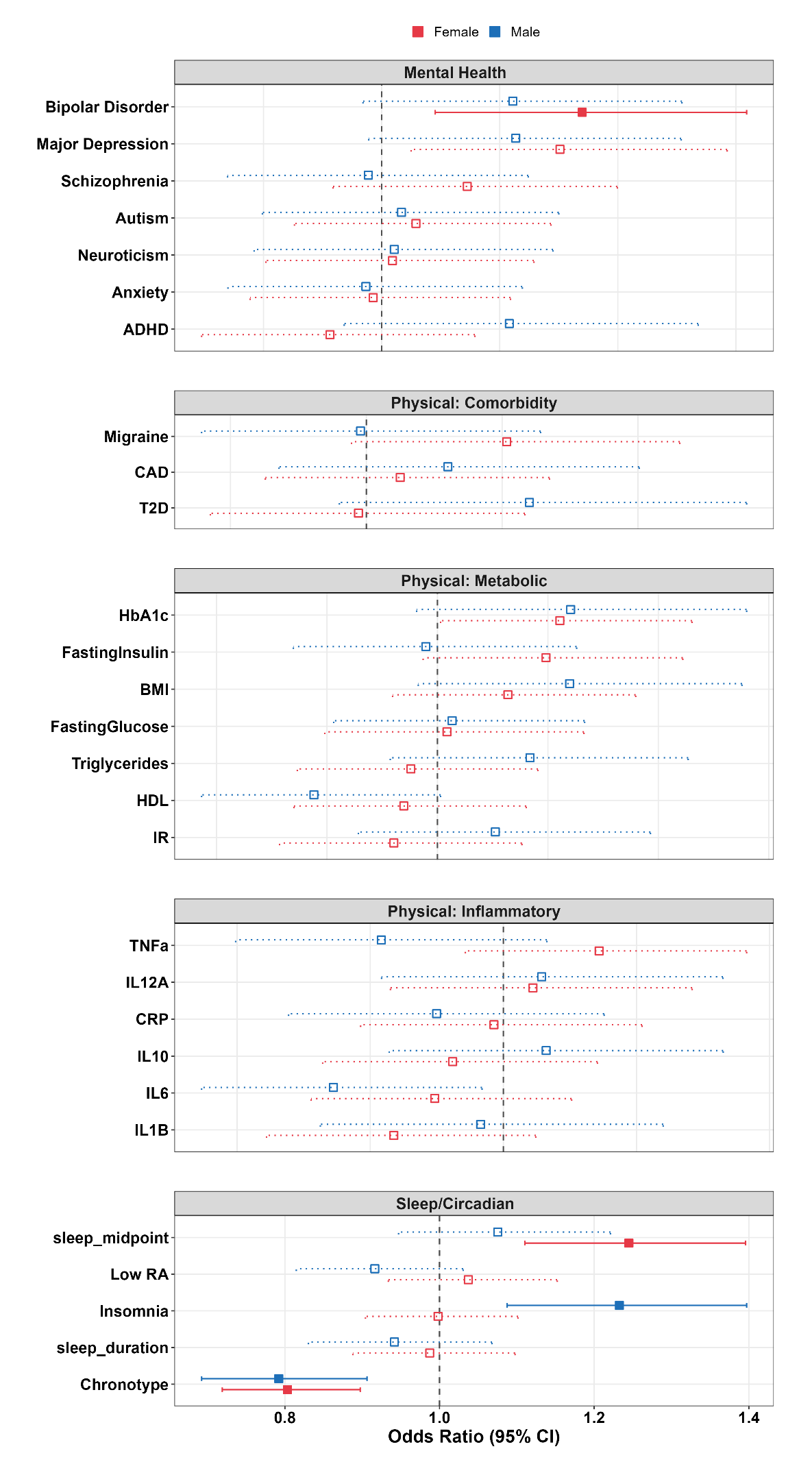

**Note:** Results shown are from separate ordinal logistic regression models for each PRS predicting the number of CRD phenotypes met (range: 0-5), fitted separately in females and males. Models were adjusted for age, twin status, and the first four genetically-inferred ancestry PCs, with cluster-robust (sandwich) standard errors clustered on family ID. Female associations are shown in red and male associations in blue. Filled squares with solid bars indicate Bonferroni-corrected significance within the relevant domain (mental health: α=0.007; physical health: α=0.003; sleep/circadian: α=0.010); hollow squares with dotted bars indicate non-significant associations.

**Abbreviations:** PRS = Polygenic Risk Score; OR = Odds Ratio; CI = Confidence Interval; ADHD = Attention Deficit Hyperactivity Disorder; HbA1c = Hemoglobin A1c; TNF-α = Tumor Necrosis Factor Alpha; IL = Interleukin; BMI = Body Mass Index; T2D = Type 2 Diabetes; CRP = C-Reactive Protein; CAD = Coronary Artery Disease; IR = Insulin Resistance; HDL = High-Density Lipoprotein Cholesterol; RA = Relative Amplitude

**Sensitivity Results 4: age-specific CRD thresholds**

Sleep timing and duration shift systematically across late adolescence and young adulthood, so thresholds derived from the full sample may over-identify circadian disturbance in younger participants and under-identify it in older participants. To test whether the findings depended on this choice, the four continuous CRD phenotypes were redefined using thresholds derived separately within participants aged 18-24 years and those aged 25 years and over, again at +1SD above the mean of the relevant age group. Seasonality and sleep inertia were unchanged, as both are defined by fixed response criteria rather than by a distribution-based cut-off. The age-specific thresholds were more stringent than the full-sample thresholds in the younger group and equal to or less stringent in the older group (Table S11).

**Table S11.** Age-specific CRD thresholds and prevalence

| **Phenotype** | **Age-specific threshold** | | | **Full-sample (primary)** | |
| --- | --- | --- | --- | --- | --- |
|  | **18-24 y** | **25+ y** | **n met (%)** | **Threshold** | **n met (%)** |
| Hypersomnia | ≥10.5 hours | ≥10 hours | 328 (12.4%) | ≥10 hours | 442 (16.7%) |
| Delayed sleep | ≥5:30am | ≥4:45am | 453 (17.1%) | ≥5:15am | 387 (14.6%) |
| Evening preference | ≥4:00am | ≥3:30am | 356 (13.4%) | ≥3:45am | 337 (12.7%) |
| Social jetlag | ≥2.5 hours | ≥2.25 hours | 358 (13.5%) | ≥2.25 hours | 405 (15.3%) |
| Seasonality^#^ | GSS ≥11 | GSS ≥11 | 59 (2.2%) | GSS ≥11 | 59 (2.2%) |
| Sleep inertia | “Very sleepy” | “Very sleepy” | 405 (15.3%) | “Very sleepy” | 405 (15.3%) |

**Note:** Percentages are of participants with available data for that phenotype. GSS = Global Seasonality Score. Seasonality was assessed only in small subsample (see the Methods section).

Redefining the thresholds shifted prevalence: Hypersomnia fell from 16.7% to 12.4% and social jetlag from 15.3% to 13.5%, whereas delayed sleep rose from 14.6% to 17.1% and evening preference from 12.7% to 13.4%. This reordered the phenotypes: delayed sleep became the most prevalent, while hypersomnia, the most prevalent under the full-sample thresholds, became the second least prevalent, and sleep inertia moved to second despite an unchanged definition (Figure S6; compare Figure 1).

**Figure S6.** Co-occurrence of circadian rhythm disturbance (CRD) phenotypes under age-specific thresholds.

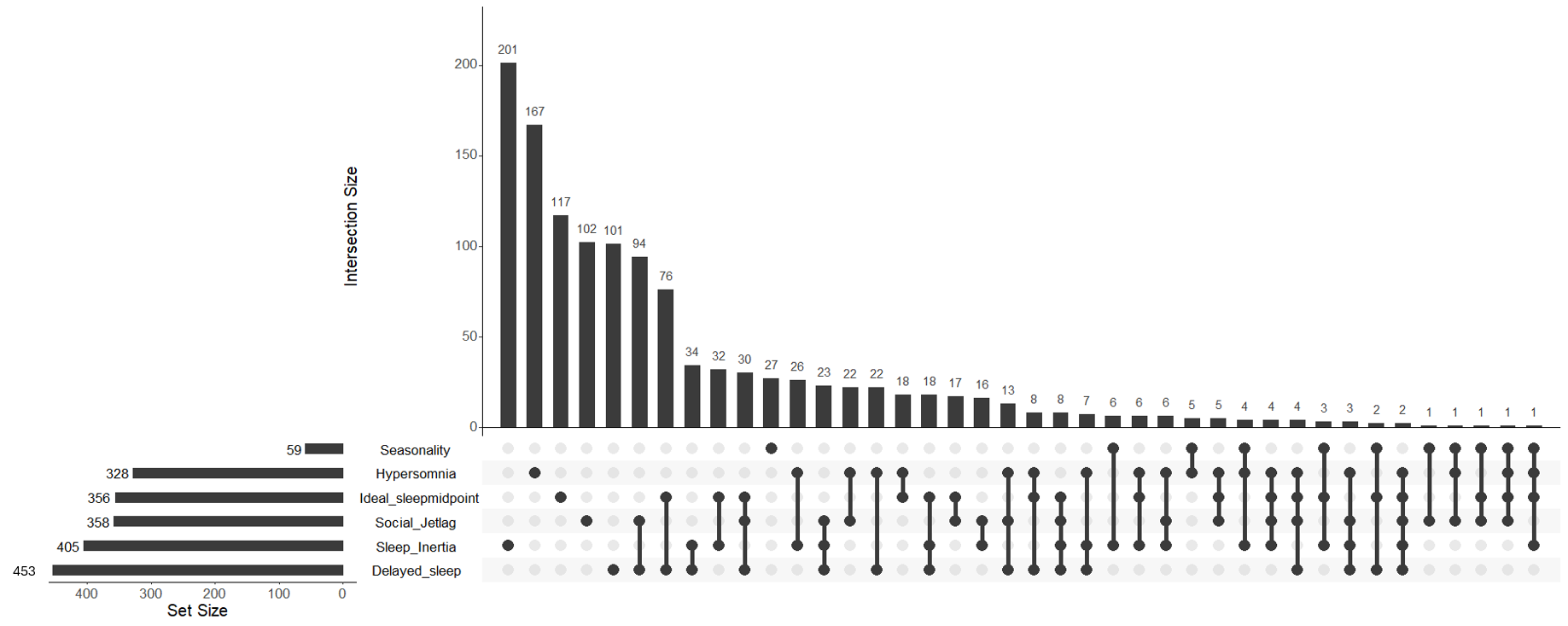

CRD-load defined by age-specific thresholds remained associated with every clinical and functional outcome examined (Table S12). Each additional CRD phenotype was associated with a 0.94-point increase in K6 psychological distress (β=0.94 [0.73, 1.16]), and with higher psychotic-like experiences (IRR=1.42), hypomanic-like experiences (IRR=1.09), somatic symptoms (IRR=1.30), psychological symptoms (IRR=1.28), days out of role (IRR=1.36), days in bed (IRR=1.52) and migraine (OR=1.47). All associations were significant at p<0.001 and survived Bonferroni correction. Estimates were close to those obtained under the pooled thresholds; the K6 coefficient, for example, was 0.94 compared with 0.96 in the primary analysis.

**Table S12.** Clinical and functional correlates of CRD-load defined by age-specific thresholds (range=0-5)

| **Outcome** | **n** | **Adjusted Estimate**  **[95% CI]** | **p** |
| --- | --- | --- | --- |
| Psychological Distress (K6)^a^ | 1692 | β=0.94 [0.73, 1.16] | <0.001* |
| Psychotic-like Experiences (CAPE)^b^ | 2651 | IRR=1.42 [1.27, 1.58] | <0.001* |
| Hypomanic-like Experiences (ASRM)^b^ | 2651 | IRR=1.09 [1.04, 1.14] | <0.001* |
| Somatic Symptoms (SPHERE)^b^ | 2651 | IRR=1.30 [1.23, 1.37] | <0.001* |
| Psychological Symptoms (SPHERE)^b^ | 2651 | IRR=1.28 [1.20, 1.37] | <0.001* |
| Days Out of Role (count) ^b^ | 1699 | IRR=1.36 [1.22, 1.53] | <0.001* |
| Days in Bed (count)^b^ | 1694 | IRR=1.52 [1.34, 1.72] | <0.001* |
| Having a Migraine ^c^ | 2651 | OR=1.47 [1.24, 1.74] | <0.001* |

**Note:** Models adjusted for age, sex, and twin status with standard errors clustered on family ID. ^a^ Using linear regression; ^b^ Using negative binomial for count-based variables due to skewed/zero-inflated counts; ^c^ Using logistic regression for binary variables. *Bonferroni-corrected significance (α=0.006). β=unstandardized coefficient; IRR=Incidence Rate Ratio; OR=Odds Ratio; CI = Confidence Interval.

PRS associations were likewise unchanged (Figure S7; SD11, SD12). Every association that survived Bonferroni correction in the primary analysis remained significant: major depression (OR=1.13 [1.04-1.23], p=0.003) and bipolar disorder (OR=1.13 [1.04-1.22], p=0.003) in the mental health domain, and chronotype (OR=0.77 [0.71-0.84], p<0.001) and later sleep midpoint (OR=1.18 [1.08-1.28], p<0.001) in the sleep/circadian domain. No physical health PRS survived correction, as in the primary analysis, and the nominal HbA1c association observed there was attenuated (OR=1.06 [0.98-1.15], p=0.12). The primary findings therefore do not depend on deriving CRD thresholds in the pooled sample.

**Figure S7**. Polygenic risk score associations with CRD-load defined by age-specific thresholds, across mental health, physical health, and sleep/circadian domains (n=2,301).

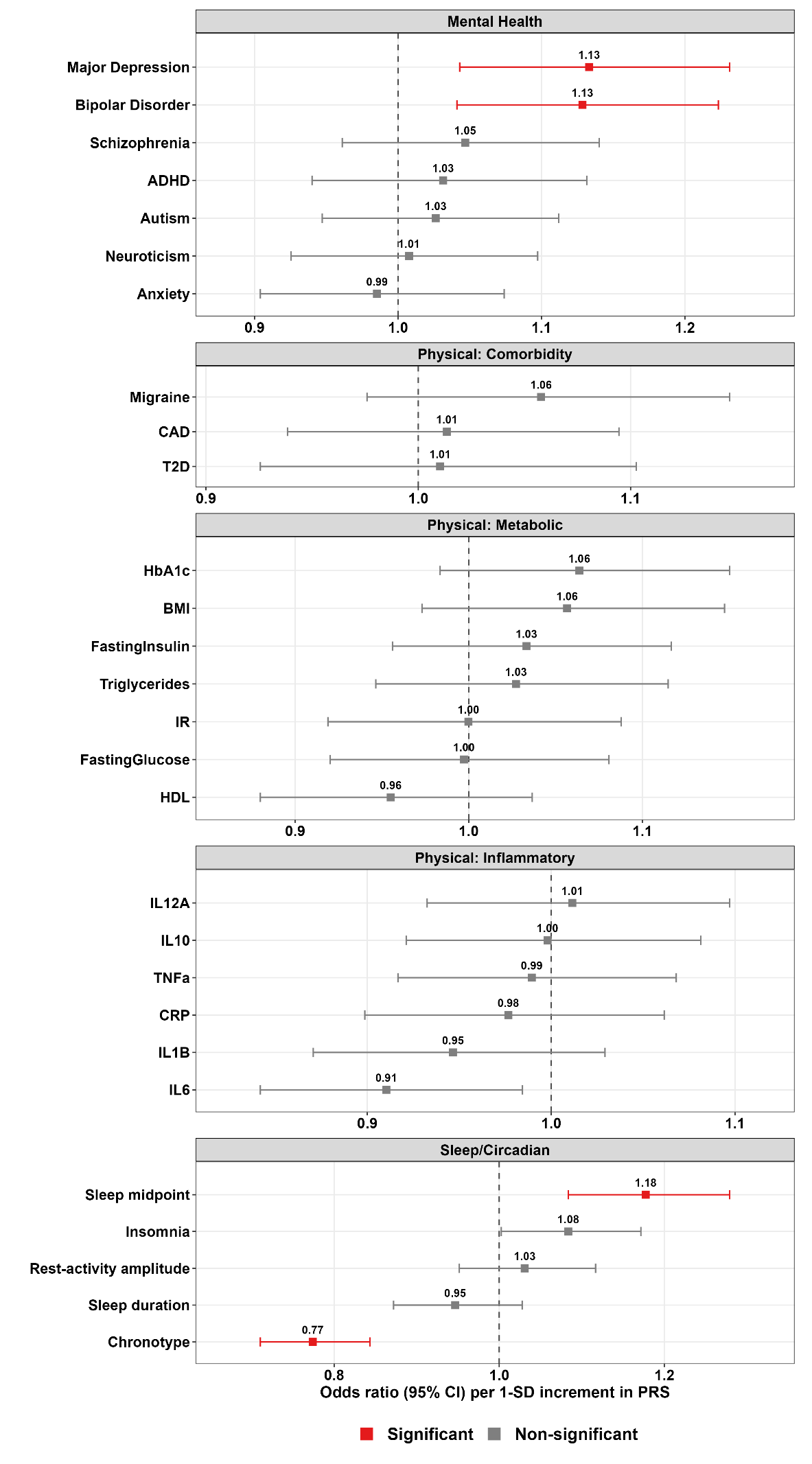

**Note:** Results shown are from separate ordinal logistic regression models for each PRS predicting the number of CRD phenotypes met under age-specific thresholds (range: 0-5), adjusted for age, sex, twin status, and the first four genetically-inferred ancestry PCs, with cluster-robust (sandwich) standard errors clustered on family ID. Red indicates Bonferroni-corrected significance within each domain (mental health: α=0.007; physical health: α=0.003; sleep/circadian: α=0.010); grey indicates non-significant associations.

**Abbreviations:** PRS = Polygenic Risk Score; OR = Odds Ratio; CI = Confidence Interval; ADHD = Attention Deficit Hyperactivity Disorder; HbA1c = Hemoglobin A1c; TNF-α = Tumor Necrosis Factor Alpha; IL = Interleukin; BMI = Body Mass Index; T2D = Type 2 Diabetes; CRP = C-Reactive Protein; CAD = Coronary Artery Disease; IR = Insulin Resistance; HDL = High-Density Lipoprotein Cholesterol; RA = Relative Amplitude

**References**

**Adams MJ, Streit F, Meng X, Awasthi S, Adey BN, Choi KW, Chundru VK, Coleman JRI, Ferwerda B, Foo JC, Gerring ZF, Giannakopoulou O, Gupta P, Hall ASM, Harder A, Howard DM, Hübel C, Kwong ASF, Levey DF, Mitchell BL, Ni G, Ota VK, Pain O, Pathak GA, Schulte EC, Shen X, Thorp JG, Walker A, Yao S, Zeng J, Zvrskovec J, Aarsland D, Actkins KEV, Adli M, Agerbo E, Aichholzer M, Aiello A, Air TM, Als TD, Andersson E, Andlauer TFM, Arolt V, Ask H, Bäckman J, Badola S, Ballard C, Banasik K, Bass NJ, Beekman ATF, Belangero S, Bigdeli TB, Binder EB, Bjerkeset O, Bjornsdottir G, Børte S, Bränn E, Braun A, Brodersen T, Brückl TM, Brunak S, Bruun MT, Burmeister M, Buspavanich P, Bybjerg-Grauholm J, Byrne EM, Cai J, Campbell A, Campbell ML, Campos AI, Castelao E, Cervilla J, Chaumette B, Chen C-Y, Chen H-C, Chen Z, Cichon S, Colodro-Conde L, Corbett A, Corfield EC, Couvy-Duchesne B, Craddock N, Dannlowski U, Davies G, de Geus EJC, Deary IJ, Degenhardt F, Dehghan A, DePaulo JR, Deuschle M, Didriksen M, Dinh KM, Direk N, Djurovic S, Docherty AR, Domschke K, Dowsett J, Drange OK, Dunn EC, Eaton W, Einarsson G, Eley TC, Elsheikh SSM, Engelmann J, Benros ME, Erikstrup C, Escott-Price V, Fabbri C, Fang Y, Finer S, Frank J, Free RC, Gallo L, Gao H, Gill M, Gilles M, Goes FS, Gordon SD, Grove J, Gudbjartsson DF, Gutierrez B, Hahn T, Hall LS, Hansen TF, Haraldsson M, Hartman CA, Havdahl A, Hayward C, Heilmann-Heimbach S, Herms S, Hickie IB, Hjalgrim H, Hjerling-Leffler J, Hoffmann P, Homuth G, Horn C, Hottenga J-J, Hougaard DM, Hovatta I, Huang QQ, Hucks D, Huider F, Hunt KA, Ialongo NS, Ising M, Isometsä E, Jansen R, Jiang Y, Jones I, Jones LA, Jonsson L, Kanai M, Karlsson R, Kasper S, Kendler KS, Kessler RC, Kloiber S, Knowles JA, Koen N, Kraft J, Kranzler HR, Krebs K, Kallak TK, Kutalik Z, Lahtela E, Lake M, Larsen MH, Lenze EJ, Lewins M, Lewis G, Li L, Lin BD, Lin K, Lind PA, Liu Y-L, MacIntyre DJ, MacKinnon DF, Maher BS, Maier W, Marshe VS, Martinez-Levy GA, Matsuda K, Mbarek H, McGuffin P, Medland SE, Meinert S, Mikkelsen C, Mikkelsen S, Milaneschi Y, Millwood IY, Molina E, Mondimore FM, Mortensen PB, Mulsant BH, Naamanka J, Najman JM, Nauck M, Nenadić I, Nielsen KR, Nolt IM, Nordentoft M, Nöthen MM, Nyegaard M, O'Donovan MC, Oddsson A, Oliveira AM, Olsen CM, Oskarsson H, Ostrowski SR, Owen MJ, Packer R, Palviainen T, Pan PM, Pato CN, Pato MT, Pedersen NL, Pedersen OB, Peyrot WJ, Potash JB, Preisig M, Preuss MH, Quiroz JA, Renteria ME, Reynolds Iii CF, Rice JP, Sakaue S, Santoro ML, Schoevers RA, Schork A, Schulze TG, Send TS, Shi J, Sigurdsson E, Singh K, Sinnamon GCB, Sirignano L, Smeland OB, Smith DJ, Sofer T, Sørensen E, Srinivasan S, Stefansson H, Stefansson K, Straub P, Su M-H, Tadic A, Teismann H, Teumer A, Thapar A, Thomson PA, Thørner LW, Topaloudi A, Tsai S-J, Tzoulaki I, Uhl G, Uitterlinden AG, Ullum H, Umbricht D, Ursano RJ, Van der Auwera S, van Hemert AM, Veluchamy A, Viktorin A, Völzke H, Walters GB, Wang X, Wani A, Weissman MM, Wellmann J, Whiteman DC, Wildman D, Willemsen G, Williams AT, Winsvold BS, Witt SH, Xiong Y, Zillich L, Zwart J-A, Twenty T, Me Research T, China Kadoorie Biobank Collaborative G, Estonian Biobank Research T, Genes, amp, Health Research T, Psychiatry HA-I, The BioBank Japan P, Program VAMV, Andreassen OA, Baune BT, Berger K, Boomsma DI, Børglum AD, Breen G, Cai N, Coon H, Copeland WE, Creese B, Cruz-Fuentes CS, Czamara D, Davis LK, Derks EM, Domenici E, Elliott P, Forstner AJ, Gawlik M, Gelernter J, Grabe HJ, Hamilton SP, Hveem K, John C, Kaprio J, Kircher T, Krebs M-O, Kuo P-H, Landén M, Lehto K, Levinson DF, Li QS, Lieb K, Loos RJF, Lu Y, Lucae S, Luykx JJ, Maes HHM, Magnusson PK, Martin HC, Martin NG, McQuillin A, Middeldorp CM, Milani L, Mors O, Müller DJ, Müller-Myhsok B, Okada Y, Oldehinkel AJ, Paciga SA, Palmer CNA, Paschou P, Penninx BWJH, Perlis RH, Peterson RE, Pistis G, Polimanti R, Porteous DJ, Posthuma D, Rabinowitz JA, Reichborn-Kjennerud T, Reif A, Rice F, Ricken R, Rietschel M, Rivera M, Rück C, Salum GA, Schaefer C, Sen S, Serretti A, Skalkidou A, Smoller JW, Stein DJ, Stein F, Stein MB, Sullivan PF, Tesli M, Thorgeirsson TE, Tiemeier H, Timpson NJ, Uddin M, Uher R, van Heel DA, Verweij KJH, Walters RG, Wassertheil-Smoller S, Wendland JR, Werge T, Zwinderman AH, Kuchenbaecker K, Wray NR, Ripke S, Lewis CM and McIntosh AM** (2025) Trans-ancestry genome-wide study of depression identifies 697 associations implicating cell types and pharmacotherapies. *Cell* **188**(3)**,** 640-652.e649. <https://doi.org/10.1016/j.cell.2024.12.002>.

**Aragam KG, Jiang T, Goel A, Kanoni S, Wolford BN, Atri DS, Weeks EM, Wang M, Hindy G, Zhou W, Grace C, Roselli C, Marston NA, Kamanu FK, Surakka I, Venegas LM, Sherliker P, Koyama S, Ishigaki K, Åsvold BO, Brown MR, Brumpton B, de Vries PS, Giannakopoulou O, Giardoglou P, Gudbjartsson DF, Güldener U, Haider SMI, Helgadottir A, Ibrahim M, Kastrati A, Kessler T, Kyriakou T, Konopka T, Li L, Ma L, Meitinger T, Mucha S, Munz M, Murgia F, Nielsen JB, Nöthen MM, Pang S, Reinberger T, Schnitzler G, Smedley D, Thorleifsson G, von Scheidt M, Ulirsch JC, Arnar DO, Burtt NP, Costanzo MC, Flannick J, Ito K, Jang DK, Kamatani Y, Khera AV, Komuro I, Kullo IJ, Lotta LA, Nelson CP, Roberts R, Thorgeirsson G, Thorsteinsdottir U, Webb TR, Baras A, Björkegren JLM, Boerwinkle E, Dedoussis G, Holm H, Hveem K, Melander O, Morrison AC, Orho-Melander M, Rallidis LS, Ruusalepp A, Sabatine MS, Stefansson K, Zalloua P, Ellinor PT, Farrall M, Danesh J, Ruff CT, Finucane HK, Hopewell JC, Clarke R, Gupta RM, Erdmann J, Samani NJ, Schunkert H, Watkins H, Willer CJ, Deloukas P, Kathiresan S and Butterworth AS** (2022) Discovery and systematic characterization of risk variants and genes for coronary artery disease in over a million participants. *Nat Genet* **54**(12)**,** 1803-1815. <https://doi.org/10.1038/s41588-022-01233-6>.

**Demontis D, Walters GB, Athanasiadis G, Walters R, Therrien K, Nielsen TT, Farajzadeh L, Voloudakis G, Bendl J, Zeng B, Zhang W, Grove J, Als TD, Duan J, Satterstrom FK, Bybjerg-Grauholm J, Bækved-Hansen M, Gudmundsson OO, Magnusson SH, Baldursson G, Davidsdottir K, Haraldsdottir GS, Agerbo E, Hoffman GE, Dalsgaard S, Martin J, Ribasés M, Boomsma DI, Soler Artigas M, Roth Mota N, Howrigan D, Medland SE, Zayats T, Rajagopal VM, Havdahl A, Doyle A, Reif A, Thapar A, Cormand B, Liao C, Burton C, Bau CHD, Rovaris DL, Sonuga-Barke E, Corfield E, Grevet EH, Larsson H, Gizer IR, Waldman I, Brikell I, Haavik J, Crosbie J, McGough J, Kuntsi J, Glessner J, Langley K, Lesch K-P, Rohde LA, Hutz MH, Klein M, Bellgrove M, Tesli M, O’Donovan MC, Andreassen OA, Leung PWL, Pan PM, Joober R, Schachar R, Loo S, Witt SH, Reichborn-Kjennerud T, Banaschewski T, Hawi Z, Daly MJ, Mors O, Nordentoft M, Mors O, Hougaard DM, Mortensen PB, Daly MJ, Faraone SV, Stefansson H, Roussos P, Franke B, Werge T, Neale BM, Stefansson K, Børglum AD, Consortium AWGotPG and i P-BC** (2023) Genome-wide analyses of ADHD identify 27 risk loci, refine the genetic architecture and implicate several cognitive domains. *Nature Genetics* **55**(2)**,** 198-208. <https://doi.org/10.1038/s41588-022-01285-8>.

**Dupuis J, Langenberg C, Prokopenko I, Saxena R, Soranzo N, Jackson AU, Wheeler E, Glazer NL, Bouatia-Naji N, Gloyn AL, Lindgren CM, Mägi R, Morris AP, Randall J, Johnson T, Elliott P, Rybin D, Thorleifsson G, Steinthorsdottir V, Henneman P, Grallert H, Dehghan A, Hottenga JJ, Franklin CS, Navarro P, Song K, Goel A, Perry JR, Egan JM, Lajunen T, Grarup N, Sparsø T, Doney A, Voight BF, Stringham HM, Li M, Kanoni S, Shrader P, Cavalcanti-Proença C, Kumari M, Qi L, Timpson NJ, Gieger C, Zabena C, Rocheleau G, Ingelsson E, An P, O'Connell J, Luan J, Elliott A, McCarroll SA, Payne F, Roccasecca RM, Pattou F, Sethupathy P, Ardlie K, Ariyurek Y, Balkau B, Barter P, Beilby JP, Ben-Shlomo Y, Benediktsson R, Bennett AJ, Bergmann S, Bochud M, Boerwinkle E, Bonnefond A, Bonnycastle LL, Borch-Johnsen K, Böttcher Y, Brunner E, Bumpstead SJ, Charpentier G, Chen YD, Chines P, Clarke R, Coin LJ, Cooper MN, Cornelis M, Crawford G, Crisponi L, Day IN, de Geus EJ, Delplanque J, Dina C, Erdos MR, Fedson AC, Fischer-Rosinsky A, Forouhi NG, Fox CS, Frants R, Franzosi MG, Galan P, Goodarzi MO, Graessler J, Groves CJ, Grundy S, Gwilliam R, Gyllensten U, Hadjadj S, Hallmans G, Hammond N, Han X, Hartikainen AL, Hassanali N, Hayward C, Heath SC, Hercberg S, Herder C, Hicks AA, Hillman DR, Hingorani AD, Hofman A, Hui J, Hung J, Isomaa B, Johnson PR, Jørgensen T, Jula A, Kaakinen M, Kaprio J, Kesaniemi YA, Kivimaki M, Knight B, Koskinen S, Kovacs P, Kyvik KO, Lathrop GM, Lawlor DA, Le Bacquer O, Lecoeur C, Li Y, Lyssenko V, Mahley R, Mangino M, Manning AK, Martínez-Larrad MT, McAteer JB, McCulloch LJ, McPherson R, Meisinger C, Melzer D, Meyre D, Mitchell BD, Morken MA, Mukherjee S, Naitza S, Narisu N, Neville MJ, Oostra BA, Orrù M, Pakyz R, Palmer CN, Paolisso G, Pattaro C, Pearson D, Peden JF, Pedersen NL, Perola M, Pfeiffer AF, Pichler I, Polasek O, Posthuma D, Potter SC, Pouta A, Province MA, Psaty BM, Rathmann W, Rayner NW, Rice K, Ripatti S, Rivadeneira F, Roden M, Rolandsson O, Sandbaek A, Sandhu M, Sanna S, Sayer AA, Scheet P, Scott LJ, Seedorf U, Sharp SJ, Shields B, Sigurethsson G, Sijbrands EJ, Silveira A, Simpson L, Singleton A, Smith NL, Sovio U, Swift A, Syddall H, Syvänen AC, Tanaka T, Thorand B, Tichet J, Tönjes A, Tuomi T, Uitterlinden AG, van Dijk KW, van Hoek M, Varma D, Visvikis-Siest S, Vitart V, Vogelzangs N, Waeber G, Wagner PJ, Walley A, Walters GB, Ward KL, Watkins H, Weedon MN, Wild SH, Willemsen G, Witteman JC, Yarnell JW, Zeggini E, Zelenika D, Zethelius B, Zhai G, Zhao JH, Zillikens MC, Borecki IB, Loos RJ, Meneton P, Magnusson PK, Nathan DM, Williams GH, Hattersley AT, Silander K, Salomaa V, Smith GD, Bornstein SR, Schwarz P, Spranger J, Karpe F, Shuldiner AR, Cooper C, Dedoussis GV, Serrano-Ríos M, Morris AD, Lind L, Palmer LJ, Hu FB, Franks PW, Ebrahim S, Marmot M, Kao WH, Pankow JS, Sampson MJ, Kuusisto J, Laakso M, Hansen T, Pedersen O, Pramstaller PP, Wichmann HE, Illig T, Rudan I, Wright AF, Stumvoll M, Campbell H, Wilson JF, Bergman RN, Buchanan TA, Collins FS, Mohlke KL, Tuomilehto J, Valle TT, Altshuler D, Rotter JI, Siscovick DS, Penninx BW, Boomsma DI, Deloukas P, Spector TD, Frayling TM, Ferrucci L, Kong A, Thorsteinsdottir U, Stefansson K, van Duijn CM, Aulchenko YS, Cao A, Scuteri A, Schlessinger D, Uda M, Ruokonen A, Jarvelin MR, Waterworth DM, Vollenweider P, Peltonen L, Mooser V, Abecasis GR, Wareham NJ, Sladek R, Froguel P, Watanabe RM, Meigs JB, Groop L, Boehnke M, McCarthy MI, Florez JC and Barroso I** (2010) New genetic loci implicated in fasting glucose homeostasis and their impact on type 2 diabetes risk. *Nat Genet* **42**(2)**,** 105-116. <https://doi.org/10.1038/ng.520>.

**Ferguson A, Lyall LM, Ward J, Strawbridge RJ, Cullen B, Graham N, Niedzwiedz CL, Johnston KJA, MacKay D, Biello SM, Pell JP, Cavanagh J, McIntosh AM, Doherty A, Bailey MES, Lyall DM, Wyse CA and Smith DJ** (2018) Genome-Wide Association Study of Circadian Rhythmicity in 71,500 UK Biobank Participants and Polygenic Association with Mood Instability. *EBioMedicine* **35,** 279-287. <https://doi.org/10.1016/j.ebiom.2018.08.004>.

**Grove J, Ripke S, Als TD, Mattheisen M, Walters RK, Won H, Pallesen J, Agerbo E, Andreassen OA, Anney R, Awashti S, Belliveau R, Bettella F, Buxbaum JD, Bybjerg-Grauholm J, Bækvad-Hansen M, Cerrato F, Chambert K, Christensen JH, Churchhouse C, Dellenvall K, Demontis D, De Rubeis S, Devlin B, Djurovic S, Dumont AL, Goldstein JI, Hansen CS, Hauberg ME, Hollegaard MV, Hope S, Howrigan DP, Huang H, Hultman CM, Klei L, Maller J, Martin J, Martin AR, Moran JL, Nyegaard M, Nærland T, Palmer DS, Palotie A, Pedersen CB, Pedersen MG, dPoterba T, Poulsen JB, Pourcain BS, Qvist P, Rehnström K, Reichenberg A, Reichert J, Robinson EB, Roeder K, Roussos P, Saemundsen E, Sandin S, Satterstrom FK, Davey Smith G, Stefansson H, Steinberg S, Stevens CR, Sullivan PF, Turley P, Walters GB, Xu X, Stefansson K, Geschwind DH, Nordentoft M, Hougaard DM, Werge T, Mors O, Mortensen PB, Neale BM, Daly MJ and Børglum AD** (2019) Identification of common genetic risk variants for autism spectrum disorder. *Nat Genet* **51**(3)**,** 431-444. <https://doi.org/10.1038/s41588-019-0344-8>.

**Hautakangas H, Winsvold BS, Ruotsalainen SE, Bjornsdottir G, Harder AVE, Kogelman LJA, Thomas LF, Noordam R, Benner C, Gormley P, Artto V, Banasik K, Bjornsdottir A, Boomsma DI, Brumpton BM, Burgdorf KS, Buring JE, Chalmer MA, de Boer I, Dichgans M, Erikstrup C, Färkkilä M, Garbrielsen ME, Ghanbari M, Hagen K, Häppölä P, Hottenga JJ, Hrafnsdottir MG, Hveem K, Johnsen MB, Kähönen M, Kristoffersen ES, Kurth T, Lehtimäki T, Lighart L, Magnusson SH, Malik R, Pedersen OB, Pelzer N, Penninx B, Ran C, Ridker PM, Rosendaal FR, Sigurdardottir GR, Skogholt AH, Sveinsson OA, Thorgeirsson TE, Ullum H, Vijfhuizen LS, Widén E, van Dijk KW, Aromaa A, Belin AC, Freilinger T, Ikram MA, Järvelin MR, Raitakari OT, Terwindt GM, Kallela M, Wessman M, Olesen J, Chasman DI, Nyholt DR, Stefánsson H, Stefansson K, van den Maagdenberg A, Hansen TF, Ripatti S, Zwart JA, Palotie A and Pirinen M** (2022) Genome-wide analysis of 102,084 migraine cases identifies 123 risk loci and subtype-specific risk alleles. *Nat Genet* **54**(2)**,** 152-160. <https://doi.org/10.1038/s41588-021-00990-0>.

**Jansen PR, Watanabe K, Stringer S, Skene N, Bryois J, Hammerschlag AR, de Leeuw CA, Benjamins JS, Muñoz-Manchado AB, Nagel M, Savage JE, Tiemeier H, White T, Tung JY, Hinds DA, Vacic V, Wang X, Sullivan PF, van der Sluis S, Polderman TJC, Smit AB, Hjerling-Leffler J, Van Someren EJW and Posthuma D** (2019) Genome-wide analysis of insomnia in 1,331,010 individuals identifies new risk loci and functional pathways. *Nat Genet* **51**(3)**,** 394-403. <https://doi.org/10.1038/s41588-018-0333-3>.

**Jones SE, Lane JM, Wood AR, van Hees VT, Tyrrell J, Beaumont RN, Jeffries AR, Dashti HS, Hillsdon M, Ruth KS, Tuke MA, Yaghootkar H, Sharp SA, Jie Y, Thompson WD, Harrison JW, Dawes A, Byrne EM, Tiemeier H, Allebrandt KV, Bowden J, Ray DW, Freathy RM, Murray A, Mazzotti DR, Gehrman PR, Lawlor DA, Frayling TM, Rutter MK, Hinds DA, Saxena R and Weedon MN** (2019) Genome-wide association analyses of chronotype in 697,828 individuals provides insights into circadian rhythms. *Nat Commun* **10**(1)**,** 343. <https://doi.org/10.1038/s41467-018-08259-7>.

**Lagou V, Mägi R, Hottenga J-J, Grallert H, Perry JRB, Bouatia-Naji N, Marullo L, Rybin D, Jansen R, Min JL, Dimas AS, Ulrich A, Zudina L, Gådin JR, Jiang L, Faggian A, Bonnefond A, Fadista J, Stathopoulou MG, Isaacs A, Willems SM, Navarro P, Tanaka T, Jackson AU, Montasser ME, O’Connell JR, Bielak LF, Webster RJ, Saxena R, Stafford JM, Pourcain BS, Timpson NJ, Salo P, Shin S-Y, Amin N, Smith AV, Li G, Verweij N, Goel A, Ford I, Johnson PCD, Johnson T, Kapur K, Thorleifsson G, Strawbridge RJ, Rasmussen-Torvik LJ, Esko T, Mihailov E, Fall T, Fraser RM, Mahajan A, Kanoni S, Giedraitis V, Kleber ME, Silbernagel G, Meyer J, Müller-Nurasyid M, Ganna A, Sarin A-P, Yengo L, Shungin D, Luan Ja, Horikoshi M, An P, Sanna S, Boettcher Y, Rayner NW, Nolte IM, Zemunik T, Iperen Ev, Kovacs P, Hastie ND, Wild SH, McLachlan S, Campbell S, Polasek O, Carlson O, Egan J, Kiess W, Willemsen G, Kuusisto J, Laakso M, Dimitriou M, Hicks AA, Rauramaa R, Bandinelli S, Thorand B, Liu Y, Miljkovic I, Lind L, Doney A, Perola M, Hingorani A, Kivimaki M, Kumari M, Bennett AJ, Groves CJ, Herder C, Koistinen HA, Kinnunen L, Faire Ud, Bakker SJL, Uusitupa M, Palmer CNA, Jukema JW, Sattar N, Pouta A, Snieder H, Boerwinkle E, Pankow JS, Magnusson PK, Krus U, Scapoli C, de Geus EJCN, Blüher M, Wolffenbuttel BHR, Province MA, Abecasis GR, Meigs JB, Hovingh GK, Lindström J, Wilson JF, Wright AF, Dedoussis GV, Bornstein SR, Schwarz PEH, Tönjes A, Winkelmann BR, Boehm BO, März W, Metspalu A, Price JF, Deloukas P, Körner A, Lakka TA, Keinanen-Kiukaanniemi SM, Saaristo TE, Bergman RN, Tuomilehto J, Wareham NJ, Langenberg C, Männistö S, Franks PW, Hayward C, Vitart V, Kaprio J, Visvikis-Siest S, Balkau B, Altshuler D, Rudan I, Stumvoll M, Campbell H, van Duijn CM, Gieger C, Illig T, Ferrucci L, Pedersen NL, Pramstaller PP, Boehnke M, Frayling TM, Shuldiner AR, Peyser PA, Kardia SLR, Palmer LJ, Penninx BW, Meneton P, Harris TB, Navis G, Harst Pvd, Smith GD, Forouhi NG, Loos RJF, Salomaa V, Soranzo N, Boomsma DI, Groop L, Tuomi T, Hofman A, Munroe PB, Gudnason V, Siscovick DS, Watkins H, Lecoeur C, Vollenweider P, Franco-Cereceda A, Eriksson P, Jarvelin M-R, Stefansson K, Hamsten A, Nicholson G, Karpe F, Dermitzakis ET, Lindgren CM, McCarthy MI, Froguel P, Kaakinen MA, Lyssenko V, Watanabe RM, Ingelsson E, Florez JC, Dupuis J, Barroso I, Morris AP, Prokopenko I, Meta-Analyses of G and Insulin-related traits C** (2021) Sex-dimorphic genetic effects and novel loci for fasting glucose and insulin variability. *Nature Communications* **12**(1)**,** 24. <https://doi.org/10.1038/s41467-020-19366-9>.

**O'Connell KS, Koromina M, van der Veen T, Boltz T, David FS, Yang JMK, Lin KH, Wang X, Coleman JRI, Mitchell BL, McGrouther CC, Rangan AV, Lind PA, Koch E, Harder A, Parker N, Bendl J, Adorjan K, Agerbo E, Albani D, Alemany S, Alliey-Rodriguez N, Als TD, Andlauer TFM, Antoniou A, Ask H, Bass N, Bauer M, Beins EC, Bigdeli TB, Pedersen CB, Boks MP, Børte S, Bosch R, Brum M, Brumpton BM, Brunkhorst-Kanaan N, Budde M, Bybjerg-Grauholm J, Byerley W, Cabana-Domínguez J, Cairns MJ, Carpiniello B, Casas M, Cervantes P, Chatzinakos C, Chen HC, Clarence T, Clarke TK, Claus I, Coombes B, Corfield EC, Cruceanu C, Cuellar-Barboza A, Czerski PM, Dafnas K, Dale AM, Dalkner N, Degenhardt F, DePaulo JR, Djurovic S, Drange OK, Escott-Price V, Fanous AH, Fellendorf FT, Ferrier IN, Forty L, Frank J, Frei O, Freimer NB, Fullard JF, Garnham J, Gizer IR, Gordon SD, Gordon-Smith K, Greenwood TA, Grove J, Guzman-Parra J, Ha TH, Hahn T, Haraldsson M, Hautzinger M, Havdahl A, Heilbronner U, Hellgren D, Herms S, Hickie IB, Hoffmann P, Holmans PA, Huang MC, Ikeda M, Jamain S, Johnson JS, Jonsson L, Kalman JL, Kamatani Y, Kennedy JL, Kim E, Kim J, Kittel-Schneider S, Knowles JA, Kogevinas M, Kranz TM, Krebs K, Kushner SA, Lavebratt C, Lawrence J, Leber M, Lee HJ, Liao C, Lucae S, Lundberg M, MacIntyre DJ, Maier W, Maihofer AX, Malaspina D, Manchia M, Maratou E, Martinsson L, Mattheisen M, McGregor NW, McInnis MG, McKay JD, Medeiros H, Meyer-Lindenberg A, Millischer V, Morris DW, Moutsatsou P, Mühleisen TW, O'Donovan C, Olsen CM, Panagiotaropoulou G, Papiol S, Pardiñas AF, Park HY, Perry A, Pfennig A, Pisanu C, Potash JB, Quested D, Rapaport MH, Regeer EJ, Rice JP, Rivera M, Schulte EC, Senner F, Shadrin A, Shilling PD, Sigurdsson E, Sindermann L, Sirignano L, Siskind D, Slaney C, Sloofman LG, Smeland OB, Smith DJ, Sobell JL, Soler Artigas M, Stein DJ, Stein F, Su MH, Sung H, Świątkowska B, Terao C, Tesfaye M, Tesli M, Thorgeirsson TE, Thorp JG, Toma C, Tondo L, Tooney PA, Tsai SJ, Tsermpini EE, Vawter MP, Vedder H, Vreeker A, Walters JTR, Winsvold BS, Witt SH, Won HH, Ye R, Young AH, Zandi PP, Zillich L, Adolfsson R, Alda M, Alfredsson L, Backlund L, Baune BT, Bellivier F, Bengesser S, Berrettini WH, Biernacka JM, Boehnke M, Børglum AD, Breen G, Carr VJ, Catts S, Cichon S, Corvin A, Craddock N, Dannlowski U, Dikeos D, Etain B, Ferentinos P, Frye M, Fullerton JM, Gawlik M, Gershon ES, Goes FS, Green MJ, Grigoroiu-Serbanescu M, Hauser J, Henskens FA, Hjerling-Leffler J, Hougaard DM, Hveem K, Iwata N, Jones I, Jones LA, Kahn RS, Kelsoe JR, Kircher T, Kirov G, Kuo PH, Landén M, Leboyer M, Li QS, Lissowska J, Lochner C, Loughland C, Luykx JJ, Martin NG, Mathews CA, Mayoral F, McElroy SL, McIntosh AM, McMahon FJ, Medland SE, Melle I, Milani L, Mitchell PB, Morken G, Mors O, Mortensen PB, Müller-Myhsok B, Myers RM, Myung W, Neale BM, Nievergelt CM, Nordentoft M, Nöthen MM, Nurnberger JI, O'Donovan MC, Oedegaard KJ, Olsson T, Owen MJ, Paciga SA, Pantelis C, Pato CN, Pato MT, Patrinos GP, Pawlak JM, Ramos-Quiroga JA, Reif A, Reininghaus EZ, Ribasés M, Rietschel M, Ripke S, Rouleau GA, Roussos P, Saito T, Schall U, Schalling M, Schofield PR, Schulze TG, Scott LJ, Scott RJ, Serretti A, Smoller JW, Squassina A, Stahl EA, Stefansson H, Stefansson K, Stordal E, Streit F, Sullivan PF, Turecki G, Vaaler AE, Vieta E, Vincent JB, Waldman ID, Weickert CS, Weickert TW, Werge T, Whiteman DC, Zwart JA, Edenberg HJ, McQuillin A, Forstner AJ, Mullins N, Di Florio A, Ophoff RA and Andreassen OA** (2025) Genomics yields biological and phenotypic insights into bipolar disorder. *Nature* **639**(8056)**,** 968-975. <https://doi.org/10.1038/s41586-024-08468-9>.

**Oliveri A, Rebernick RJ, Kuppa A, Pant A, Chen Y, Du X, Cushing KC, Bell HN, Raut C, Prabhu P, Chen VL, Halligan BD and Speliotes EK** (2024) Comprehensive genetic study of the insulin resistance marker TG:HDL-C in the UK Biobank. *Nat Genet* **56**(2)**,** 212-221. <https://doi.org/10.1038/s41588-023-01625-2>.

**Rosenthal NE, Sack DA, Gillin JC, Lewy AJ, Goodwin FK, Davenport Y, Mueller PS, Newsome DA and Wehr TA** (1984) Seasonal affective disorder. A description of the syndrome and preliminary findings with light therapy. *Arch Gen Psychiatry* **41**(1)**,** 72-80. <https://doi.org/10.1001/archpsyc.1984.01790120076010>.

**Said S, Pazoki R, Karhunen V, Võsa U, Ligthart S, Bodinier B, Koskeridis F, Welsh P, Alizadeh BZ, Chasman DI, Sattar N, Chadeau-Hyam M, Evangelou E, Jarvelin M-R, Elliott P, Tzoulaki I and Dehghan A** (2022) Genetic analysis of over half a million people characterises C-reactive protein loci. *Nature Communications* **13**(1)**,** 2198. <https://doi.org/10.1038/s41467-022-29650-5>.

**Schwaba T, Sullivan MLC, Akingbuwa WA, Ilves K, Tanksley PT, Williams CM, Dragostinov Y, Liao W, Ackerman LS and Fealy JC** (2025) Robust inference and widespread genetic correlates from a large-scale genetic association study of human personality. *Biorxiv*.

**Sinnott-Armstrong N, Tanigawa Y, Amar D, Mars N, Benner C, Aguirre M, Venkataraman GR, Wainberg M, Ollila HM, Kiiskinen T, Havulinna AS, Pirruccello JP, Qian J, Shcherbina A, Rodriguez F, Assimes TL, Agarwala V, Tibshirani R, Hastie T, Ripatti S, Pritchard JK, Daly MJ, Rivas MA and FinnGen** (2021) Genetics of 35 blood and urine biomarkers in the UK Biobank. *Nature Genetics* **53**(2)**,** 185-194. <https://doi.org/10.1038/s41588-020-00757-z>.

**Strom NI, Verhulst B, Bacanu S-A, Cheesman R, Purves KL, Gedik H, Mitchell BL, Kwong AS, Faucon AB, Singh K, Medland S, Colodro-Conde L, Krebs K, Hoffmann P, Herms S, Gehlen J, Ripke S, Awasthi S, Palviainen T, Tasanko EM, Peterson RE, Adkins DE, Shabalin AA, Adams MJ, Iveson MH, Campbell A, Thomas LF, Winsvold BS, Drange OK, Børte S, ter Kuile AR, Naamanka J, Nguyen T-H, Meier SM, Corfield EC, Hannigan L, Levey DF, Czamara D, Weber H, Choi KW, Pistis G, Couvy-Duchesne B, Van der Auwera S, Teumer A, Karlsson R, Garcia-Argibay M, Lee D, Wang R, Bjerkeset O, Stordal E, Bäckman J, Salum GA, Zai CC, Kennedy JL, Zai G, Tiwari AK, Heilmann-Heimbach S, Schmidt B, Kaprio J, Kennedy MM, Boden J, Havdahl A, Middeldorp CM, Lopes FL, Akula N, McMahon FJ, Binder EB, Fehm L, Ströhle A, Castelao E, Tiemeier H, Stein DJ, Whiteman D, Olsen C, Fuller Z, Wang X, Wray NR, Byrne EM, Lewis G, Timpson NJ, Davis LK, Hickie IB, Gillespie NA, Milani L, Schumacher J, Woldbye DP, Forstner AJ, Nöthen MM, Hovatta I, Horwood J, Copeland WE, Maes HH, McIntosh AM, Andreassen OA, Zwart J-A, Mors O, Børglum AD, Mortensen PB, Ask H, Reichborn-Kjennerud T, Najman JM, Stein MB, Gelernter J, Milaneschi Y, Penninx BW, Boomsma DI, Maron E, Erhardt-Lehmann A, Rück C, Kircher TT, Melzig CA, Alpers GW, Arolt V, Domschke K, Smoller JW, Preisig M, Martin NG, Lupton MK, Luik AI, Reif A, Grabe HJ, Larsson H, Magnusson PK, Oldehinkel AJ, Hartman CA, Breen G, Docherty AR, Coon H, Conrad R, Lehto K, Deckert J, Eley TC, Mattheisen M, Hettema JM, Veterans Affairs Million Veteran P, FinnGen and andMe Research T** (2026) Genome-wide association study of major anxiety disorders in 122,341 European-ancestry cases identifies 58 loci and highlights GABAergic signaling. *Nature Genetics* **58**(2)**,** 275-288. <https://doi.org/10.1038/s41588-025-02485-8>.

**Sun BB, Chiou J, Traylor M, Benner C, Hsu Y-H, Richardson TG, Surendran P, Mahajan A, Robins C, Vasquez-Grinnell SG, Hou L, Kvikstad EM, Burren OS, Davitte J, Ferber KL, Gillies CE, Hedman ÅK, Hu S, Lin T, Mikkilineni R, Pendergrass RK, Pickering C, Prins B, Baird D, Chen C-Y, Ward LD, Deaton AM, Welsh S, Willis CM, Lehner N, Arnold M, Wörheide MA, Suhre K, Kastenmüller G, Sethi A, Cule M, Raj A, Kang HM, Burkitt-Gray L, Melamud E, Black MH, Fauman EB, Howson JMM, Kang HM, McCarthy MI, Nioi P, Petrovski S, Scott RA, Smith EN, Szalma S, Waterworth DM, Mitnaul LJ, Szustakowski JD, Gibson BW, Miller MR, Whelan CD, Alnylam Human G, AstraZeneca Genomics I, Biogen Biobank T, Bristol Myers S, Genentech Human G, GlaxoSmithKline Genomic S, Pfizer Integrative B, Population Analytics of Janssen Data S and Regeneron Genetics C** (2023) Plasma proteomic associations with genetics and health in the UK Biobank. *Nature* **622**(7982)**,** 329-338. <https://doi.org/10.1038/s41586-023-06592-6>.

**Suzuki K, Hatzikotoulas K, Southam L, Taylor HJ, Yin X, Lorenz KM, Mandla R, Huerta-Chagoya A, Melloni GEM, Kanoni S, Rayner NW, Bocher O, Arruda AL, Sonehara K, Namba S, Lee SSK, Preuss MH, Petty LE, Schroeder P, Vanderwerff B, Kals M, Bragg F, Lin K, Guo X, Zhang W, Yao J, Kim YJ, Graff M, Takeuchi F, Nano J, Lamri A, Nakatochi M, Moon S, Scott RA, Cook JP, Lee J-J, Pan I, Taliun D, Parra EJ, Chai J-F, Bielak LF, Tabara Y, Hai Y, Thorleifsson G, Grarup N, Sofer T, Wuttke M, Sarnowski C, Gieger C, Nousome D, Trompet S, Kwak S-H, Long J, Sun M, Tong L, Chen W-M, Nongmaithem SS, Noordam R, Lim VJY, Tam CHT, Joo YY, Chen C-H, Raffield LM, Prins BP, Nicolas A, Yanek LR, Chen G, Brody JA, Kabagambe E, An P, Xiang AH, Choi HS, Cade BE, Tan J, Broadaway KA, Williamson A, Kamali Z, Cui J, Thangam M, Adair LS, Adeyemo A, Aguilar-Salinas CA, Ahluwalia TS, Anand SS, Bertoni A, Bork-Jensen J, Brandslund I, Buchanan TA, Burant CF, Butterworth AS, Canouil M, Chan JCN, Chang L-C, Chee M-L, Chen J, Chen S-H, Chen Y-T, Chen Z, Chuang L-M, Cushman M, Danesh J, Das SK, de Silva HJ, Dedoussis G, Dimitrov L, Doumatey AP, Du S, Duan Q, Eckardt K-U, Emery LS, Evans DS, Evans MK, Fischer K, Floyd JS, Ford I, Franco OH, Frayling TM, Freedman BI, Genter P, Gerstein HC, Giedraitis V, González-Villalpando C, González-Villalpando ME, Gordon-Larsen P, Gross M, Guare LA, Hackinger S, Hakaste L, Han S, Hattersley AT, Herder C, Horikoshi M, Howard A-G, Hsueh W, Huang M, Huang W, Hung Y-J, Hwang MY, Hwu C-M, Ichihara S, Ikram MA, Ingelsson M, Islam MT, Isono M, Jang H-M, Jasmine F, Jiang G, Jonas JB, Jørgensen T, Kamanu FK, Kandeel FR, Kasturiratne A, Katsuya T, Kaur V, Kawaguchi T, Keaton JM, Kho AN, Khor C-C, Kibriya MG, Kim D-H, Kronenberg F, Kuusisto J, Läll K, Lange LA, Lee KM, Lee M-S, Lee NR, Leong A, Li L, Li Y, Li-Gao R, Ligthart S, Lindgren CM, Linneberg A, Liu C-T, Liu J, Locke AE, Louie T, Luan Ja, Luk AO, Luo X, Lv J, Lynch JA, Lyssenko V, Maeda S, Mamakou V, Mansuri SR, Matsuda K, Meitinger T, Melander O, Metspalu A, Mo H, Morris AD, Moura FA, Nadler JL, Nalls MA, Nayak U, Ntalla I, Okada Y, Orozco L, Patel SR, Patil S, Pei P, Pereira MA, Peters A, Pirie FJ, Polikowsky HG, Porneala B, Prasad G, Rasmussen-Torvik LJ, Reiner AP, Roden M, Rohde R, Roll K, Sabanayagam C, Sandow K, Sankareswaran A, Sattar N, Schönherr S, Shahriar M, Shen B, Shi J, Shin DM, Shojima N, Smith JA, So WY, Stančáková A, Steinthorsdottir V, Stilp AM, Strauch K, Taylor KD, Thorand B, Thorsteinsdottir U, Tomlinson B, Tran TC, Tsai F-J, Tuomilehto J, Tusie-Luna T, Udler MS, Valladares-Salgado A, van Dam RM, van Klinken JB, Varma R, Wacher-Rodarte N, Wheeler E, Wickremasinghe AR, van Dijk KW, Witte DR, Yajnik CS, Yamamoto K, Yamamoto K, Yoon K, Yu C, Yuan J-M, Yusuf S, Zawistowski M, Zhang L, Zheng W, Kanona S, van Heel DA, Raffel LJ, Igase M, Ipp E, Redline S, Cho YS, Lind L, Province MA, Fornage M, Hanis CL, Ingelsson E, Zonderman AB, Psaty BM, Wang Y-X, Rotimi CN, Becker DM, Matsuda F, Liu Y, Yokota M, Kardia SLR, Peyser PA, Pankow JS, Engert JC, Bonnefond A, Froguel P, Wilson JG, Sheu WHH, Wu J-Y, Hayes MG, Ma RCW, Wong T-Y, Mook-Kanamori DO, Program VAMV, Japan AGDI, Biobank Japan P, Penn Medicine B, Regeneron Genetics C, Genes, Health Research T, e MC, International Consortium of Blood P, Meta-Analyses of G and Insulin-Related Traits C** (2024) Genetic drivers of heterogeneity in type 2 diabetes pathophysiology. *Nature* **627**(8003)**,** 347-357. <https://doi.org/10.1038/s41586-024-07019-6>.

**Trubetskoy V, Pardiñas AF, Qi T, Panagiotaropoulou G, Awasthi S, Bigdeli TB, Bryois J, Chen CY, Dennison CA, Hall LS, Lam M, Watanabe K, Frei O, Ge T, Harwood JC, Koopmans F, Magnusson S, Richards AL, Sidorenko J, Wu Y, Zeng J, Grove J, Kim M, Li Z, Voloudakis G, Zhang W, Adams M, Agartz I, Atkinson EG, Agerbo E, Al Eissa M, Albus M, Alexander M, Alizadeh BZ, Alptekin K, Als TD, Amin F, Arolt V, Arrojo M, Athanasiu L, Azevedo MH, Bacanu SA, Bass NJ, Begemann M, Belliveau RA, Bene J, Benyamin B, Bergen SE, Blasi G, Bobes J, Bonassi S, Braun A, Bressan RA, Bromet EJ, Bruggeman R, Buckley PF, Buckner RL, Bybjerg-Grauholm J, Cahn W, Cairns MJ, Calkins ME, Carr VJ, Castle D, Catts SV, Chambert KD, Chan RCK, Chaumette B, Cheng W, Cheung EFC, Chong SA, Cohen D, Consoli A, Cordeiro Q, Costas J, Curtis C, Davidson M, Davis KL, de Haan L, Degenhardt F, DeLisi LE, Demontis D, Dickerson F, Dikeos D, Dinan T, Djurovic S, Duan J, Ducci G, Dudbridge F, Eriksson JG, Fañanás L, Faraone SV, Fiorentino A, Forstner A, Frank J, Freimer NB, Fromer M, Frustaci A, Gadelha A, Genovese G, Gershon ES, Giannitelli M, Giegling I, Giusti-Rodríguez P, Godard S, Goldstein JI, González Peñas J, González-Pinto A, Gopal S, Gratten J, Green MF, Greenwood TA, Guillin O, Gülöksüz S, Gur RE, Gur RC, Gutiérrez B, Hahn E, Hakonarson H, Haroutunian V, Hartmann AM, Harvey C, Hayward C, Henskens FA, Herms S, Hoffmann P, Howrigan DP, Ikeda M, Iyegbe C, Joa I, Julià A, Kähler AK, Kam-Thong T, Kamatani Y, Karachanak-Yankova S, Kebir O, Keller MC, Kelly BJ, Khrunin A, Kim SW, Klovins J, Kondratiev N, Konte B, Kraft J, Kubo M, Kučinskas V, Kučinskiene ZA, Kusumawardhani A, Kuzelova-Ptackova H, Landi S, Lazzeroni LC, Lee PH, Legge SE, Lehrer DS, Lencer R, Lerer B, Li M, Lieberman J, Light GA, Limborska S, Liu CM, Lönnqvist J, Loughland CM, Lubinski J, Luykx JJ, Lynham A, Macek M, Jr., Mackinnon A, Magnusson PKE, Maher BS, Maier W, Malaspina D, Mallet J, Marder SR, Marsal S, Martin AR, Martorell L, Mattheisen M, McCarley RW, McDonald C, McGrath JJ, Medeiros H, Meier S, Melegh B, Melle I, Mesholam-Gately RI, Metspalu A, Michie PT, Milani L, Milanova V, Mitjans M, Molden E, Molina E, Molto MD, Mondelli V, Moreno C, Morley CP, Muntané G, Murphy KC, Myin-Germeys I, Nenadić I, Nestadt G, Nikitina-Zake L, Noto C, Nuechterlein KH, O'Brien NL, O'Neill FA, Oh SY, Olincy A, Ota VK, Pantelis C, Papadimitriou GN, Parellada M, Paunio T, Pellegrino R, Periyasamy S, Perkins DO, Pfuhlmann B, Pietiläinen O, Pimm J, Porteous D, Powell J, Quattrone D, Quested D, Radant AD, Rampino A, Rapaport MH, Rautanen A, Reichenberg A, Roe C, Roffman JL, Roth J, Rothermundt M, Rutten BPF, Saker-Delye S, Salomaa V, Sanjuan J, Santoro ML, Savitz A, Schall U, Scott RJ, Seidman LJ, Sharp SI, Shi J, Siever LJ, Sigurdsson E, Sim K, Skarabis N, Slominsky P, So HC, Sobell JL, Söderman E, Stain HJ, Steen NE, Steixner-Kumar AA, Stögmann E, Stone WS, Straub RE, Streit F, Strengman E, Stroup TS, Subramaniam M, Sugar CA, Suvisaari J, Svrakic DM, Swerdlow NR, Szatkiewicz JP, Ta TMT, Takahashi A, Terao C, Thibaut F, Toncheva D, Tooney PA, Torretta S, Tosato S, Tura GB, Turetsky BI, Üçok A, Vaaler A, van Amelsvoort T, van Winkel R, Veijola J, Waddington J, Walter H, Waterreus A, Webb BT, Weiser M, Williams NM, Witt SH, Wormley BK, Wu JQ, Xu Z, Yolken R, Zai CC, Zhou W, Zhu F, Zimprich F, Atbaşoğlu EC, Ayub M, Benner C, Bertolino A, Black DW, Bray NJ, Breen G, Buccola NG, Byerley WF, Chen WJ, Cloninger CR, Crespo-Facorro B, Donohoe G, Freedman R, Galletly C, Gandal MJ, Gennarelli M, Hougaard DM, Hwu HG, Jablensky AV, McCarroll SA, Moran JL, Mors O, Mortensen PB, Müller-Myhsok B, Neil AL, Nordentoft M, Pato MT, Petryshen TL, Pirinen M, Pulver AE, Schulze TG, Silverman JM, Smoller JW, Stahl EA, Tsuang DW, Vilella E, Wang SH, Xu S, Adolfsson R, Arango C, Baune BT, Belangero SI, Børglum AD, Braff D, Bramon E, Buxbaum JD, Campion D, Cervilla JA, Cichon S, Collier DA, Corvin A, Curtis D, Forti MD, Domenici E, Ehrenreich H, Escott-Price V, Esko T, Fanous AH, Gareeva A, Gawlik M, Gejman PV, Gill M, Glatt SJ, Golimbet V, Hong KS, Hultman CM, Hyman SE, Iwata N, Jönsson EG, Kahn RS, Kennedy JL, Khusnutdinova E, Kirov G, Knowles JA, Krebs MO, Laurent-Levinson C, Lee J, Lencz T, Levinson DF, Li QS, Liu J, Malhotra AK, Malhotra D, McIntosh A, McQuillin A, Menezes PR, Morgan VA, Morris DW, Mowry BJ, Murray RM, Nimgaonkar V, Nöthen MM, Ophoff RA, Paciga SA, Palotie A, Pato CN, Qin S, Rietschel M, Riley BP, Rivera M, Rujescu D, Saka MC, Sanders AR, Schwab SG, Serretti A, Sham PC, Shi Y, St Clair D, Stefánsson H, Stefansson K, Tsuang MT, van Os J, Vawter MP, Weinberger DR, Werge T, Wildenauer DB, Yu X, Yue W, Holmans PA, Pocklington AJ, Roussos P, Vassos E, Verhage M, Visscher PM, Yang J, Posthuma D, Andreassen OA, Kendler KS, Owen MJ, Wray NR, Daly MJ, Huang H, Neale BM, Sullivan PF, Ripke S, Walters JTR and O'Donovan MC** (2022) Mapping genomic loci implicates genes and synaptic biology in schizophrenia. *Nature* **604**(7906)**,** 502-508. <https://doi.org/10.1038/s41586-022-04434-5>.

**Yengo L, Sidorenko J, Kemper KE, Zheng Z, Wood AR, Weedon MN, Frayling TM, Hirschhorn J, Yang J, Visscher PM and Consortium tG** (2018) Meta-analysis of genome-wide association studies for height and body mass index in ∼700000 individuals of European ancestry. *Human Molecular Genetics* **27**(20)**,** 3641-3649. <https://doi.org/10.1093/hmg/ddy271>.
